# Primary Central Nervous System Lymphoma Tumor Biopsies Show Heterogeneity in Gene Expression Profiles and Genetic Subtypes

**DOI:** 10.1101/2024.11.11.24316310

**Authors:** Yuan Xiao Zhu, Jasper CH Wong, Talal Hilal, Alanna Maguire, Jon Ocal, Katie Zellner, Xianfeng Chen, Brian K Link, Thomas M Habermann, Matthew J Maurer, James R Cerhan, Patrick B Johnston, Andrew L Feldman, David W. Scott, Allison Rosenthal, Lisa Rimsza

## Abstract

**Background:** Primary central nervous system lymphoma (PCNSL) is clinically challenging due to its location and small biopsy size, leading to a lack of comprehensive molecular and biologic description. We previously demonstrated that 91% of PCNSL belong to the activated B-cell-like (ABC) molecular subtype of diffuse large B-cell lymphoma (DLBCL).

**Methods:** Here we investigated the expression of 739 cancer related genes in HIV(-) patients in 25 ABC-PCNSL and 43 ABC-systemic DLBCL, all tumors were EBV(-).

**Results:** We found 135 genes which were identified as differentially expressed between these ABC-PCNSL and ABC-systemic DLBCL (p<0.05). The ABC-PCNSL showed higher gene expression in several cancer-related gene sets including genes related to Hedgehog, DNA damage repair, Wnt and MAPK signaling. Whole exome sequencing showed distinct genetic features of PCNSL compared to DLBCL, CXCR4 mutations in particular, that have been associated with ibrutinib resistance. In a focused analysis, PCNSL and DLBCL cases that fall into the “MCD” genetic subtype showed substantial overlap. These data provide detailed information about unique characteristics of PCNSL in HIV(-) patients as compared to comparable DLBCL subtypes.

## Introduction

Primary central nervous system large B-cell lymphoma (PCNSL) is a mature B-cell lymphoma with large cell cytology, predominantly of the activated B-cell-like (ABC) subtype of diffuse large B-cell lymphomas (DLBCL) ^1^ that presents and remains confined to the central nervous system ^2^. PCNSL may occur in immunocompromised patients with human immunodeficiency virus (HIV) and express Epstein-Barr virus associated mRNAs (EBER). However, most cases arise in patients that are HIV(-) and whose tumors lack EBER ^3^. PCNSL is included in the global category of large B-cell lymphomas of immune-privileged sites, alongside cases occurring in the testis or vitreoretina, with the rationale that these locations may share common mechanisms of lymphomagenesis ^2^.

Despite a favorable initial response to chemotherapy and radiation, the prognosis of PCNSL is inferior to that of DLBCL presenting outside the CNS (systemic DLBCL). The incidence of PCNSL has increased in older adults (>65-70 years) over the last four decades while improvement in survival has not ^4^, likely due to the difficulty of safely administering intensive CNS-penetrating chemotherapy in that age group ^5^. In a nationwide population-based study, the 2-year and 5-year overall survival among patients >60 years old was 44% and 28%, respectively ^6^. Identification of potential therapeutic targets and targeted agents that could be administered to older adults is therefore of clinical interest.

Herein, we compared PCNSL to systemic DLBCL using gene expression profiling (GEP), mutation and copy number analysis (CNA), correlated with patient survival data and drug sensitivity assays to preliminarily identify altered cellular pathways that may be amenable to therapeutic targeting. We focused on PCNSL (cases with secondary CNS involvement were excluded) from patients without HIV infection, EBV(-) tumors, and ABC subtype determined by gold standard molecular methods, to create a uniform cohort of patients and comparisons.

## Materials and Methods

### Patient samples

Previously, a cohort of PCNSL and systemic DLBCL patient diagnostic samples with annotated clinical outcomes was obtained from the Molecular Epidemiology Resource (MER) ^7^. Cases with secondary involvement of the CNS were excluded. For rigorous comparison, the current work focused exclusively on HIV(-) patients and EBV(-) PCNSL tumors. GEP analysis was restricted to cases (N=25) from our previous work that had both adequate remaining material and expression patterns that were molecularly confirmed as Activated B cell subtype (ABC), in order to remove the known differences in gene expression between the ABC and Germinal center B-cell like (GCB) subtypes of large cell lymphoma ^1^. However, three additional PCNSL cases from the previous work that had typed as GCB were included in the sequencing analyses for completeness (total of N = 28 PCNSL), and with the added rationale that the ABC/GCB GEP subtyping assay and algorithm were trained on systemic DLBCL and may not be the most relevant classification system for evaluating PCNSL genetics. The systemic DLBCL cases (N=43) were likewise HIV(-), EBV(-), ABC subtype, and were retrieved from the MER cohort. Hematoxylin and eosin (H&E) stained slides were reviewed to determine tumor content. Cases with less than 60% tumor content were enriched by macro-dissection. This study was conducted in compliance with IRB approved protocols and the Declaration of Helsinki (PR16-000507 and PR2207-02).

### Cell lines

Two human PCNSL cell lines, TK and HKBML, were purchased from the JCRB cell bank and RIKEN BRC cell bank in Japan, respectively. Both cell lines have been reported and used in previous PCNSL research ^8,9^ and were the only 2 confirmed PCNSL lines commercially available. TK cells were cultured in RPMI1640 medium supplemented with 10% fetal calf serum (FCS) and HKBML cells were maintained in HamF12 with 15% FCS. A human diffuse large B-cell lymphoma cell line (HBL-1) was gifted from Dr. Riccardo Dalla-Favera’s lab (Columbia University, New York, NY) and maintained in IMDM with 10% FCS. Two additional human B-cell lymphoma cell lines (OCI-Ly3 and TMD8) were gifted from Dr. Louis Staudt’s lab at the National Cancer Institute (Bethesda, MD) and maintained in RPMI with 10% FCS.

### DNA and RNA extraction

DNA and RNA were extracted with the AllPrep DNA/RNA FFPE kit (Qiagen, USA) from at least 2 x 10 µm formalin-fixed paraffin-embedded (FFPE) unstained sections. RNA was quantified using a NanoDrop (Thermo-Scientific, USA). DNA was quantified using the High Sensitivity dsDNA Qubit kit (Invitrogen, USA).

### NanoString and gene expression analysis

The Cell of Origin (COO) based on GEP had been determined in our previous work using the Lymph2Cx assay ^1,10^. For the current work, RNA input of 300ng per patient sample or 100ng per cell line was used for a modified PanCancer Pathways (739 target genes plus 40 housekeepers) panel from NanoString (Seattle, WA), which included 606 pathway genes for 13 canonical pathways, 124 cancer driver genes, and 9 BCL2 family genes. GEP assays were performed on the NanoString nCounter system (Seattle, WA). RNA from an immortalized DLBCL cell line (DB cells, ATCC) was included as a batch control in each run for quality control and normalization (supplemental figure 1). The collected RCC data were first evaluated using the NanoString nSolver 4.0 Analysis software for quality control, followed by technical normalization using synthetic controls, and biological normalization via housekeeping genes. The data collected were then analyzed with an Advanced Analysis software plugin (version 2.0, R-based statistical tool) for nSolver 4.0 to detect and visualize differentially expressed genes, perform unsupervised hierarchical clustering, and assess pathway score changes in different groups of samples.

### Whole exome sequencing (WES), copy number, and EBV analysis

For sequencing analyses, 25 ABC-PCNSL cases, 3 GCB-PCNSL cases, and 5 cell lines were sequenced using the Agilent SureSelect Human All Exon v6 + UTR kit at Canada’s Michael Smith Genome Sciences Center at BC Cancer, Vancouver, Canada. Libraries were sequenced on an Illumina high-throughput lane (150bp paired-ends). Reads were aligned to GRCh37 reference genome using BWA-MEM (v.0.7.17) ^11^. High-confidence simple somatic variants (SSMs) were determined using SLMS-3, a voting approach of four variant callers (Strelka2, Lofreq, Mutect2, and SAGE), wherein consensus variants were selected if three out of four variant callers identified them ^12–14^. SAGE can be found at https://github.com/hartwigmedical/hmftools/tree/master/sage. The full pipeline can be found on https://github.com/LCR-BCCRC/lcr-modules (slms3). The genetic subtype of each sample was determined based on the mutational data and classified using the well-known LymphGen algorithm, as previously described ^15^.

Potential copy number alterations (CNAs) were identified using PureCN (v.2.0.1) ^16^, which calculated and corrected for tumor ploidy and purity. High confidence CNAs were determined using GISTIC v2.0.23 ^17^ with a q-value cut-off of 0.1. Samples showing a tumor purity of 0.4 or less were excluded from copy number analyses due to high levels of noise (Supplemental data 1). Manual review of the CDKN2A locus was performed using CNVkit (v.0.9.9) ^18^. Observations were subsequently cross-referenced with the loci of known oncogenes and tumor suppressors implicated in DLBCL. GISTIC plots were generated using Oviz-Bio’s visualization suite ^19^.

Mutational and copy number profiles were compared to published ABC-systemic DLBCL WESlibraries generated by Agilent SureSelect v2 ^20^ and Agilent SureSelect v5 ^21,22^. Mutational calls were restricted to regions shared by all capture kits. Publicly available datasets were processed using PureCN (v.2.0.1).

Regions that contained high repeats, segmental duplications, and centromeric and telomeric regions were excluded from copy number analyses.

To determine EBV positivity, the GATK (v.4.1.8.1) pipeline PathSeq was used to map any unmapped reads in WES libraries to a reference EBV genome with a threshold of 0.0002% reads. We leveraged other WES libraries that used the same capture kit with experimentally confirmed EBV status to further threshold the cohort.

### Immunoblotting assays

Western blots used 30mg of protein subjected to sodium dodecyl sulfate polyacrylamide gel electrophoresis (SDS-PAGE) gels followed by transfer to PVDF membranes. Membranes were probed with primary antibodies overnight, washed, and incubated with horseradish peroxidase (HRP)- conjugated-secondary antibodies. Detection was performed by the Enhanced Chemical Luminescence (ECL) method. Anti-AKT, anti-pAKT (S473), anti-PERK (137F5), anti-ERK (Thr202/Y204) and anti b-catenin antibodies were from Cell Signaling Technology (Danvers, MA), and the mouse anti-β-actin antibody was from GenScript (Piscataway, NJ).

### Drug treatment and cell viability assay

PCNSL and ABC-systemic DLBCL cell lines were treated with methotrexate (MTX) and temozolomide (TMZ), 2 commonly used chemotherapy drugs as well as novel agents and drugs inhibiting highly expressed pathways in PCNSL, including Bruton’s tyrosine kinase (BTK) (ibrutinib), AKT (afuresertib), which is a key component of the PI3K pathway, BCL2 (venetoclax), immunomodulatory drugs (pomalidomide), and fibroblastic growth factor receptor (FGFR) (BGJ398). All drugs were purchased from Selleck Chemicals LLC (Houston, TX). Cell viability and cell growth were measured by MTS or MTT cell proliferation assay kits according to the manufacturer’s instructions (Abcam, Waltham, MA). Experiments were performed in triplicate and repeated at least twice.

### Clinical survival analyses

Kaplan-Meier survival analyses based on either GEP features or genetic subgroups were performed using R library ggsur rvp lot (https://cloud.r-project.org/package=survminer). Overall survival was defined from the time of diagnosis to death from any cause.

## RESULTS

### PCNSL shows higher gene expression in Hedgehog, DNA damage repair, Wnt and MAPK associated signaling compared to systemic DLBCL

We first compared the expression of 739 cancer-related genes in 25 cases of ABC-PCNSL with 43 cases of ABC-systemic DLBCL (which had sufficient remaining material from our previous publication). The heatmap of normalized expression data indicated that most of the ABC-PCNSL cohort (P) had a distinct transcriptional landscape from that of the ABC-systemic DLBCL cohort (non-PCNSL systemic, abbreviated “NS”) (Figure 1A). Of the 739 genes, 135 (18.3%) showed significant differential expression between these two groups (p<0.05, adj p<0.05, supplemental data 2 and Figure 1B). Analysis showed significant upregulation of KMT2D, MCL1, NTRK2 and MAPK10, and downregulation of LAMA5 and FLNA (Figure 1C and Supplemental data 2), in the ABC-PCNSL compared with the ABC-systemic DLBCL cases. Using pathway score analysis (provided in the analysis software), which condenses each samples’ pathway gene expression profiles into small sets of pathway scores (increasing score corresponds to mostly increasing expression), we identified that ABC-PCNSL had higher pathway scores in signaling related to Hedgehog, DNA damage repair, Wnt, and MAPK, and had lower pathway scores in the gene sets associated with cell cycle-apoptosis, Notch and Jak-stat signaling and driver genes (Figure 1C).

**Figure 1.**
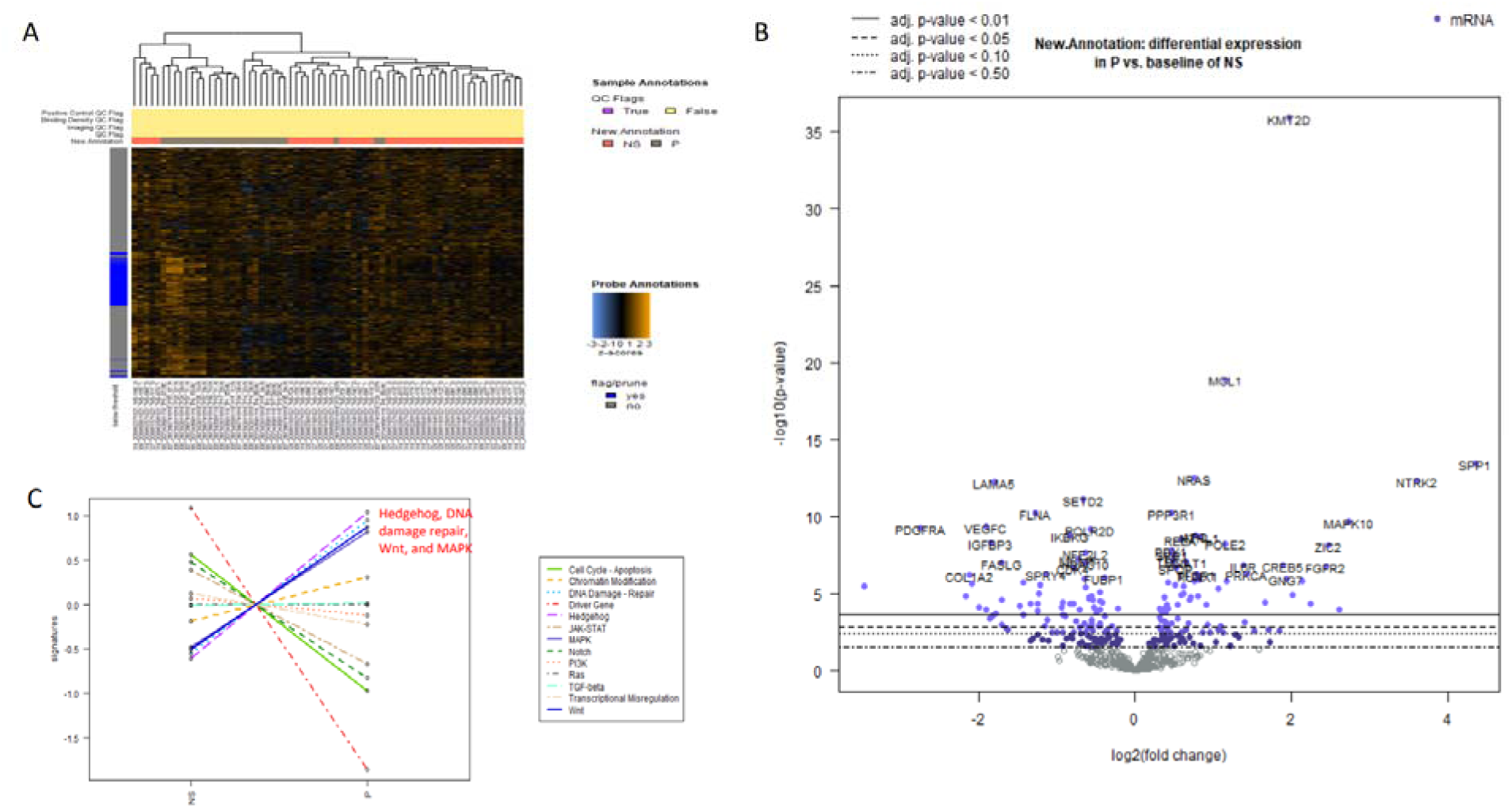
Differentially expressed genes between PCNSL (P) and non-PCNSL/systemic DLBCL (NS) were identified. (A) Heatmap overview of normalized expression data of P and NS on the PanCancer Pathways panel. The heatmap was scaled to give all genes equal variance and was generated via unsupervised clustering. Orange indicates high expression; blue indicates low expression. This plot is meant to provide a high-level exploratory view of the data. (B) A volcano plot displaying each gene’s log10(p-value) and log2 fold change with the selected covariate. Genes with high statistical significance lie at the top of the plot above the horizontal lines, and genes with high differential expression fall to either side. Horizontal lines indicate the various False Discovery Rates (FDR) or appropriate p-value thresholds if there is no adjustment to the p-values. Genes are colored if the resulting p-value is below the given FDR or p-value threshold. The 40 most statistically significant genes are labeled in the plot. (C) Covariate plots showing pathway score analysis of P and NS indicated a higher gene expression in Hedgehog, DNA damage repair, Wnt and MAPK associated signaling compared to ABC- systemic DLBCL.

To determine whether some of the differentially expressed genes between ABC-PCNSL and ABC- systemic DLBCL could originate from low purity related to background CNS tissues (Supplemental data 1), we compared the differentially expressed gene list with the 474 CNS-enriched genes identified from a published report of gene expression in normal CNS compared to malignant B cells ^23^. We found that 5 CNS-enriched genes from Montesinos-Rongen et al. were on our list, including FGFR2, HDAC10, JAG2, MAPK8IP1 and SKP1 (Supplemental data 3). We further compared our list with another 475 highly expressed genes in the human brain from a published dataset (Human Protein Atlas) and identified 3 genes that were on our list (Supplemental data 3), including FGFR2, GNG7, and ZIC2. To explore these results, immunohistochemistry staining of FGFR2 protein was performed. While FGFR2 protein was expressed in kidney tubules (positive control), little to no protein staining was seen in the normal brain and normal tonsil. In contrast, FGFR2 protein expression in PCNSL was mainly detected in the surrounding brain tissue and stromal- rich areas, indicating an alteration of the brain tissue surrounding the tumor cells (Supplemental Figure 2).

### PCNSL separated into two groups based on gene expression data with differing survival

In an exploratory analysis using unsupervised hierarchical clustering of all 28 PCNSL samples, we found two clusters of cases that we termed PCNSL group 1 (P1, n=9) and PCNSL group 2 (P2, n= 19) (Figure 2A). The P1 and P2 samples were from different runs and contained both ABC and GCB types (one GCB sample in P2, two in P1) confirming this was not a batch effect and suggesting that this grouping may override the cell-of-origin (Figure 2B). The P2 group had more samples with high tumor purity (based on copy number data) (Figure 2B, 44%) than the P1 group (23.5%), but one subcluster in the P1 group showed a similar percentage of high-purity samples (40%) to P2. Further analysis indicated that P2 had higher pathway scores across most of the cancer related pathways (mainly PI3K, MAPK and RAS), but DNA damage repair pathways were lower (Figure 2C-D). We then correlated the two transcriptional subtypes with survival data (Supplemental data 4). We used a Cox proportional hazards model (Proportional Hazards Regression Model), including age as a covariate, to rule out the impact of age on survival. Patients with a P2 subtype showed a significantly shorter overall survival, p=0.003 (Figure 3A). Analysis of differentially expressed genes between P2 and P1 identified 171 genes of interest (p<0.05, Supplemental data 5), with the top 10 associated with gene sets of chromatin modification, Ras, cell cycle-apoptosis, PI3K, TGF-beta, MAPK and JAK-STAT (Figure 3B).

**Figure 2.**
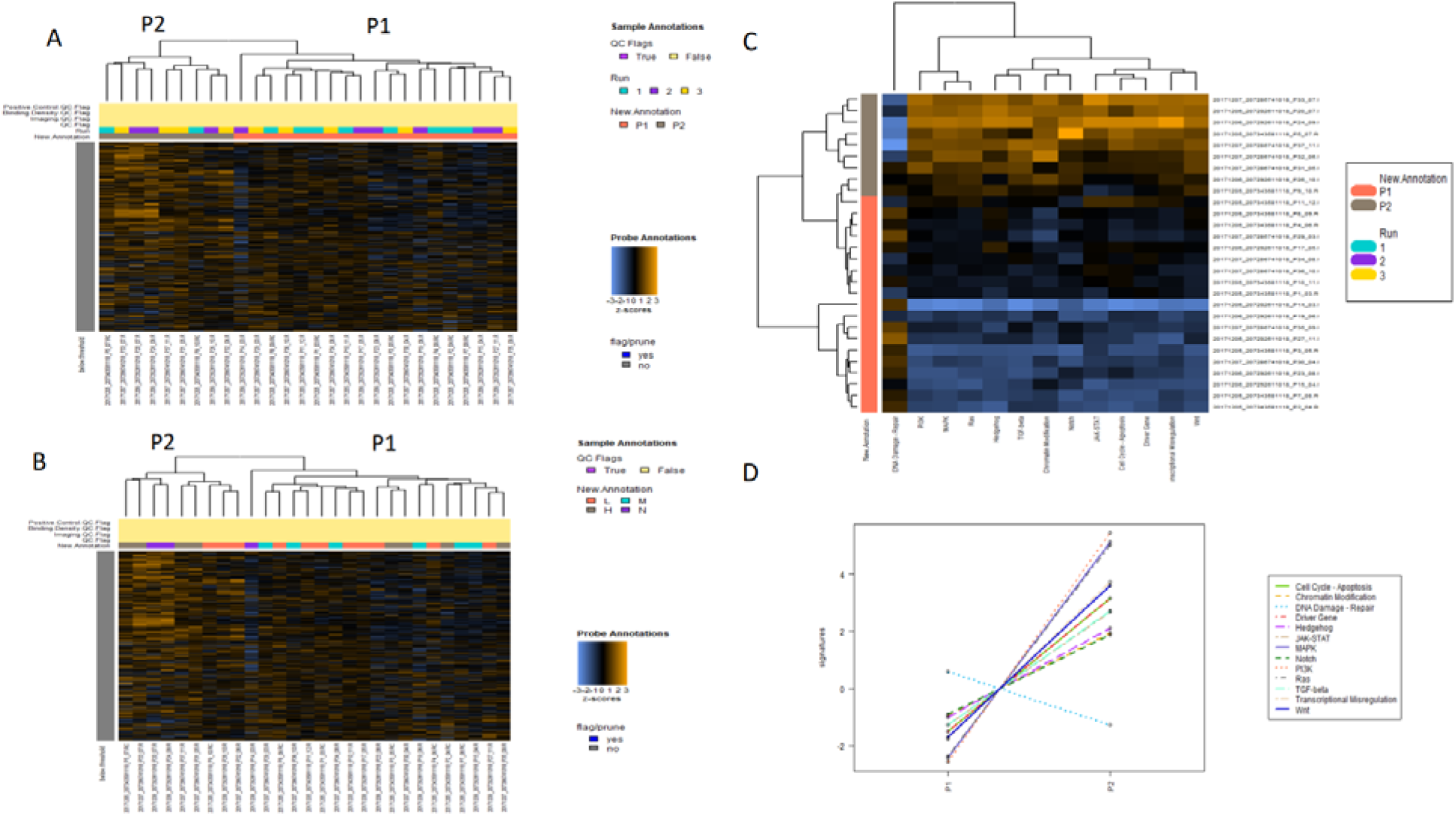
Two transcriptional subgroups were identified in PCNSL samples. (A-B) These analyses show 2 heatmaps of normalized data from PCNSL revealing two transcriptional clusters (P1 and P2) using unsupervised hierarchical clustering. The samples from different runs (A) and with different tumor purities (B) were marked demonstrating that these clusters were not the result of different batches or purity-related artifacts. In (B) High/H: tumor purity ≥ 0.6; Medium/M: tumor purity between 0.3 and 0.6; Low/L: tumor purity ≤ 0.3; N: no information about purity. (C) Heatmap of pathway scores across all PCNSL samples indicated that the P2 group had higher “pathway scores” across most of the cancer related pathways as annotated on the X-axis while the P1 group had higher expression of genes related to DNA damage. (D) Covariate plots comparing the pathway scores between P2 and P1 identified top pathway score changes, including high scores in PI3K, MAPK, RAS signaling and low scores in DNA damage signaling in the P2 group.

**Figure 3.**
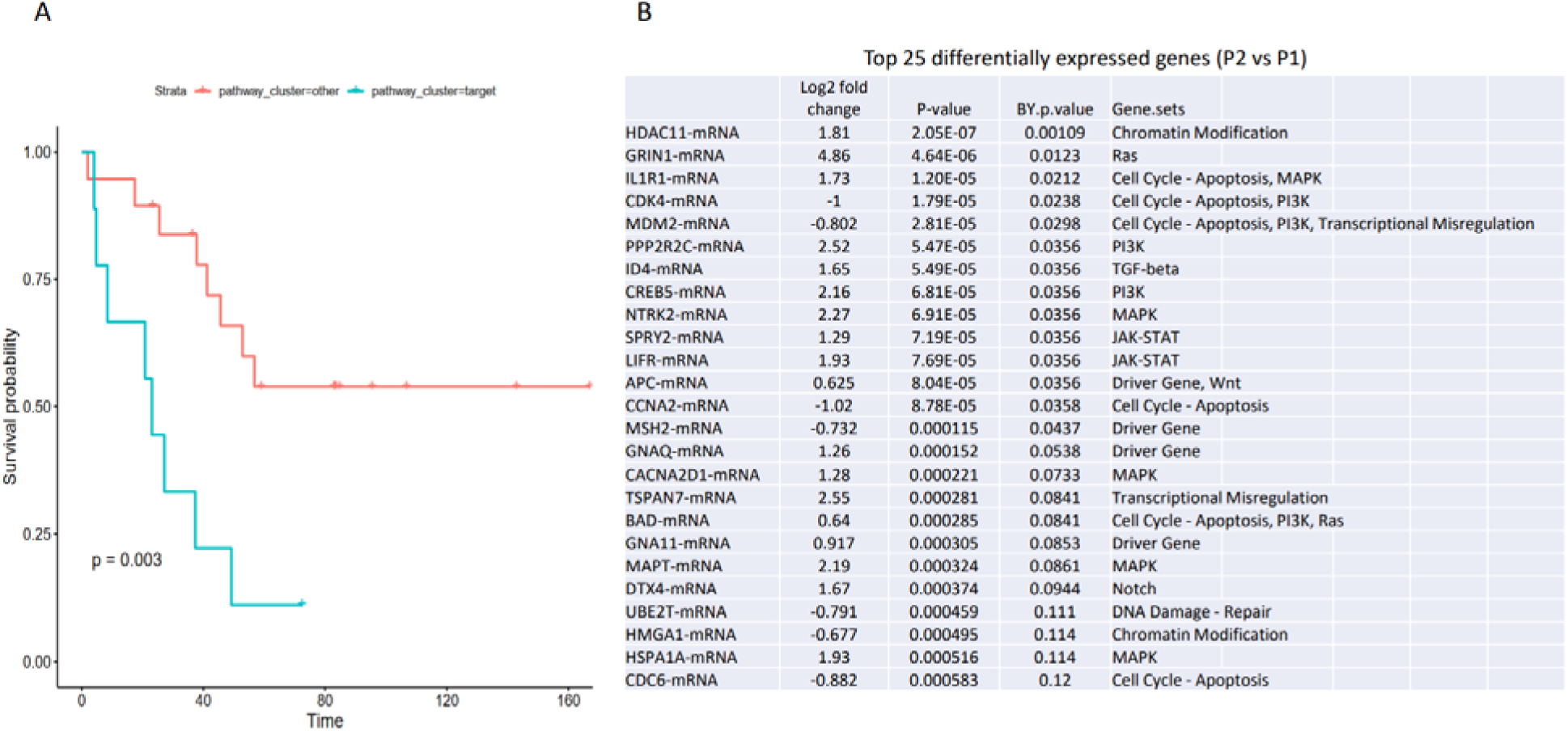
The patients in P2 have a short clinical overall survival. (A) Kaplan-Meier survival analysis of patient data from two transcriptional subgroups of PCNSL demonstrated a shorter survival time for patients in the P2 subgroup compared with the P1 subgroup (p=0.003). (B) The top 25 genes with the greatest differential expression between P1 and P2 are listed in descending order of significance.

### Mutational landscape, confirmation of EBV status, and copy number alterations in PCNSL

We performed WES to determine the mutational landscape and genetic subtypes of the PCNSL cases, as originally described for DLBCL known as the LymphGen subtype ^15^. For completeness and contrast, both the 25 ABC-PCNSL and 3 GCB-PCNSL cases were included in the initial analysis. As expected for a cohort with an ABC predominance, the majority of PCNSL patient samples had mutations in MYD88^L265P^ (18/29; 62.1%), PIM1 (17/29; 58.6%), and CD79B (14/29; 48.3%), representative of the “MCD” LymphGen subclass. MYD88^L265P^ is a key mutation defining ABC- systemic DLBCL and, by inference, ABC-PCNSL (Figure 4A). Indeed, using the mutational data with the LymphGen algorithm online tool, 59% of PCNSL samples were identified as “MCD”, 17% as “MCD-composite” (MCD in combination with other subtypes), and 24% as “Other” (not classifiable into a particular category) (Figure 4B)^15^. Consistent with previous descriptions of PCNSL, the most frequent gene mutations affected the NF-kB pathway including MYD88^L265P^ (62%), CD79B (48%), CARD11 (23%), TBL1XR1 (27%), and NFKBIZ 3’UTR mutations (14%).

**Figure 4.**
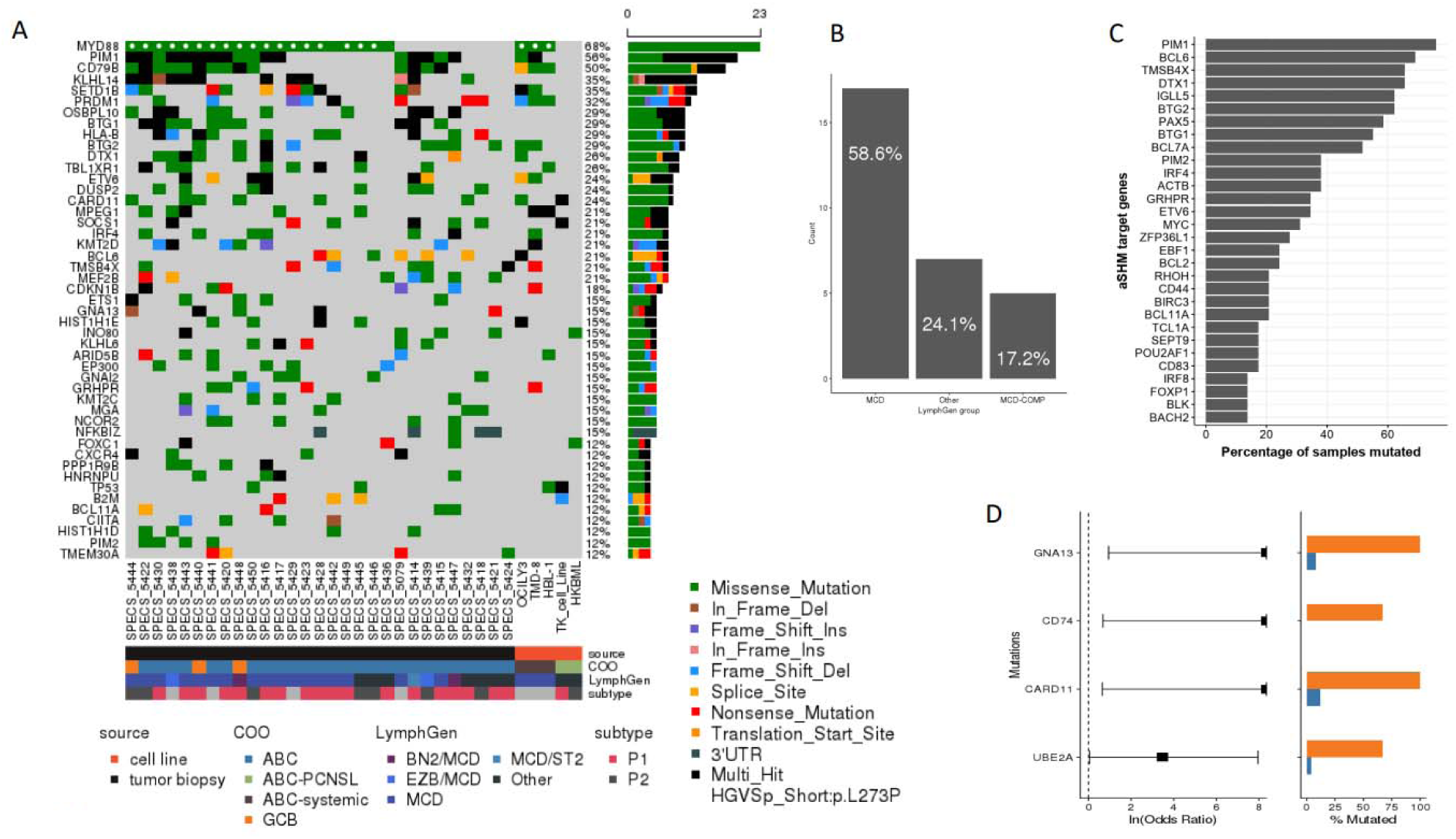
WES analysis revealed genetic heterogeneity in PCNSL. (A-B) Oncoplot showing the mutational landscape of PCNSL patient cases and cell lines derived from ABC-PCNSL and ABC-systemic DLBCL. The tracks below the Oncoplot show additional pathological features, including COO, LymphGen subgroup assignment, and PCNSL expression subtype. The majority (59%) of ABC-PCNSL patient samples were identified as “MCD” by the mutational algorithm, 17% as “MCD-composite” (MCD with another subtype), and 24% as “Other” (not classifiable into a particular subtype). (C) Bar plot showing the percentage of PCNSL with mutations in common aSHM regions that have been described in DLBCL. (D) Fisher’s exact tests were conducted between ABC-PCNSL (blue) and GCB-PCNSL (orange). The forest plot and corresponding bar plot revealed mutations in GNA13, CD74, CARD11, and UBE2A as significantly (q < 0.1) associated with the uncommon GCB-PCNSL (similar to GCB-systemic DLBCL).

Additional common mutations known to be associated with the MCD-DLBCL subtype, including KLHL14 (40%), SETD1B (33%), HLA-B (33%), BTG1 (33%), OSBPL10 (33%), PRDM1 (27%), ETV6 (20%), IRF4 (20%), and MPEG1 (17%) were found in the PCNSL cases ^15^. We also found DUSP2 (27%) mutations, which have been associated with higher FOXP3 and PD-L1 positive cell populations in PCNSL ^24^. No association of LymphGen subtypes with clinical survival was observed in our cohort, likely due to low case numbers (data not shown).

Since the WES libraries covered the non-coding space, we also examined the 3’UTR and 5’UTR areas of regions that have been described as targets of aberrant somatic hypermutation (aSHM) ^25,26^. In the PCNSL cases, the predominant genes that demonstrated frequent mutations were PIM1 (75%), BCL6 (69%), DTX1 (66%), TMSB4X (66%), IGLL5 (62%), BTG2 (62%), PAX5 (59%), BTG1 (55%), and BCL7A (52%) (Figure 4C). Notably, mutations were centered around the transcription start site (TSS) of MYC (31%) and BCL2 (24%) (Supplemental Figure 3).

When relating these mutational patterns to the transcriptional subtypes we observed in the GEP analysis, no genetic lesions were enriched in either the P1 or P2 group (Supplemental Figure 4). Interestingly, within our initial cohort of PCNSL cases, three samples had been classified as GCB. As an additional confirmatory check, we compared the mutational profile of the GCB-PCNSL against ABC- PCNSL. Similar to the published literature for GCB-systemic DLBCL, we observed significant enrichment (q < 0.1, Fisher’s exact test) in GNA13 (3/3, 100%), CARD11 (3/3, 100%), and UBE2A (2/3, 67%) mutations in GCB-PCNSL, while ABC-PCNSL harbored fewer GNA13 mutations (2/26, 8%), CARD11 mutations (3/26, 12%), and UBE2A mutations (1/26, 4%) cases (Figure 4D). When comparing regions that have been annotated as common aSHM sites in systemic DLBCL, we observed unique mutations in the CD74 locus around the TSS within 2 of the 3 GCB-PCNSL cases (67%). These results need to be verified in a larger cohort of the uncommon GCB-PCNSL cases.

To address the fact that EBV(+) PCNSL tumors are biologically distinct from EBV(-) PCNSL, we confirmed the EBV(-) status of our cases by querying unmapped reads in the WES libraries that mapped to the EBV genome. Only one case showed some read support for EBV, though it was just below the threshold for consideration (Supplemental Figure 5). All other PCNSL samples in our cohort showed no reads aligning to the EBV genome.

Using PureCN v2.0.1 to evaluate ploidy and copy number alterations in PCNSL, we determined the tumor purity and correlated the results with ploidy of the PCNSL tumors from the WES libraries, demonstrating 31% (9/29) of samples displayed polyploidy. We next confined the data set to 16 samples that had a tumor purity of at least 40% for further analyses with GISTIC v.2.0.23. Arm-level amplifications affected chromosomes 7p (44%), 12p (31%), 12q (31%), and 18q (44%), while arm-level deletions affected chromosome 6q (56%). Notably, focal amplifications were observed in 31% (5/16) of tumors at 2p13.3, which contains the gene encoding the splicing regulator PCBP-1 (PCBP1). Amplifications at 14q24.1 (ZFP36L1), 3q26.32 (TBL1XR1), and 1p13.1 (PDE4B) were also observed in 43% (7/16), 31% (5/16), and 31% (5/16) of cases, respectively. Recurrent focal deletions at 9p21.3 (CDKN2A) and 19p13.11 (MEF2B) were observed in 56% (9/16) and 31% (5/16) of the PCNSL tumors, respectively (Supplemental Figure 6).

As in previous publications, the three ABC-systemic DLBCL lines (OCI-Ly3, TMD-8, HBL-1) were classified as “MCD” and harbored mutations representative of the “MCD” subclass (MYD88^L265P^, CD79B, and PRDM1) ^15,27^. In comparison, both ABC-PCNSL cell lines (TK and HKBML) lacked aberrations in the canonical MCD genes and were classified as “Other” by LymphGen (Figure 4A and Supplemental Figure 7). Notably, the PCNSL TK line displayed a mutation in CARD11, which has been associated with increased resistance to ibrutinib ^28^. One of the ABC-systemic DLBCL lines, OCI-Ly3, also harbored CARD11 mutations as previously reported.

### Subset analyses of PCNSL and systemic-DLBCL mutational data and copy number data in the cases with ABC expression profile and MCD genetic features

In order to compare the mutational landscape of our PCNSL cases to ABC-systemic DLBCL, we broadened the systemic DLBCL data set by gathering 395 exome data sets of ABC-systemic DLBCL from published sources ^20–22^. We first compared our ABC-PCNSL cases to all ABC-systemic DLBCL. Overall, ABC-PCNSL showed a significantly (P < 0.01, t-test) higher tumor mutation burden (TMB) compared to ABC- systemic DLBCL with a median TMB of 2.86 mutations/MB compared to 2.31 mutations/MB in ABC- systemic DLBCL.

Since the genetic subtypes are thought to be more homogenous than the GEP-defined subgroups, and the majority of PCNSL cases were identified as the “MCD” LymphGen subgroup, in a more rigorous subset analysis, we compared PCNSL to MCD-classified ABC-systemic DLBCL (MCD-ABC-systemic DLBCL) as the most biologically comparable group. Compared to the 112 MCD-ABC-systemic DLBCL, the TMB of PCNSL was similar to that of the MCD-ABC-systemic DLBCL with a median of 2.62 mutations/MB (Figure 5A).

**Figure 5.**
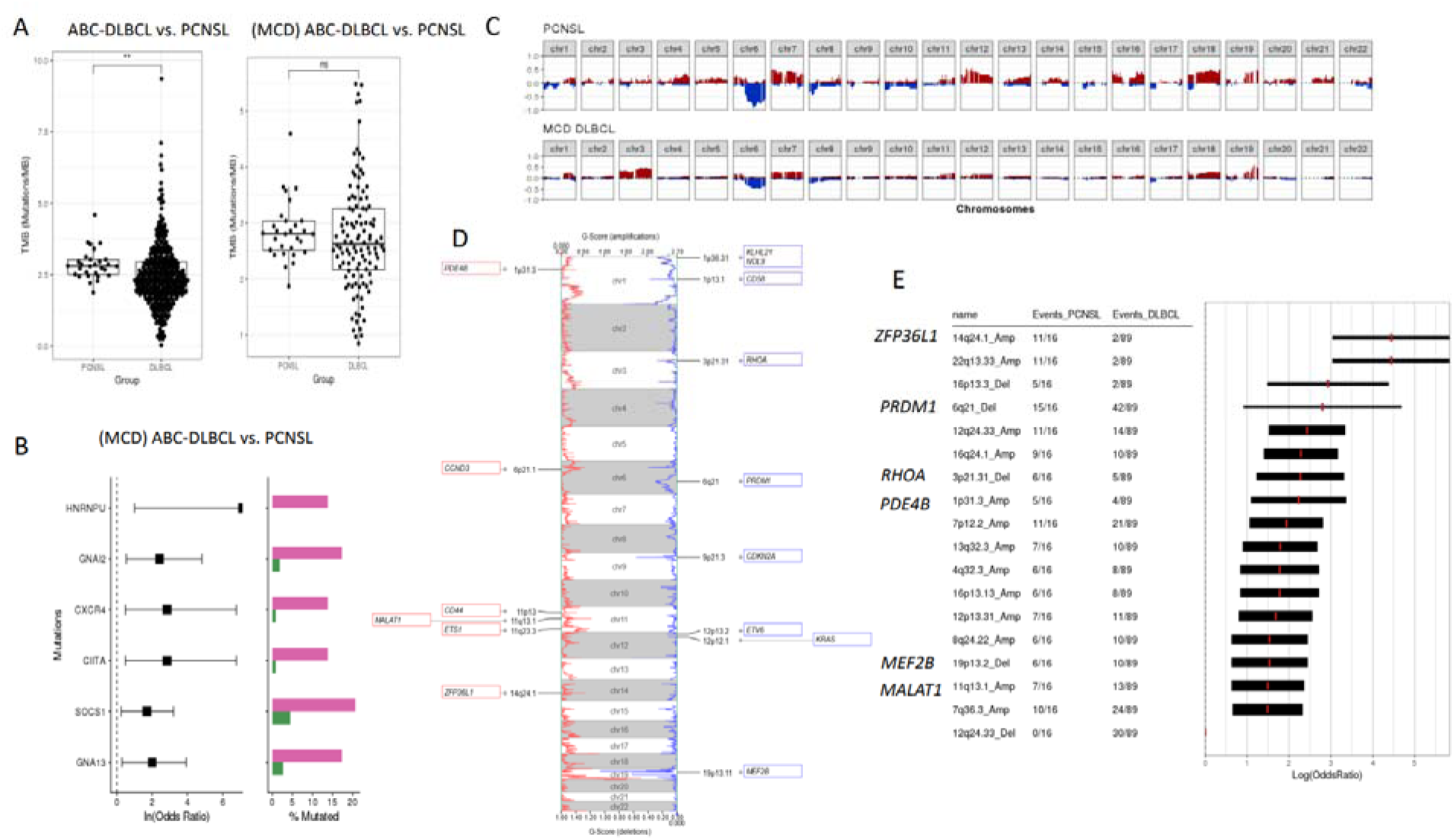
PCNSL demonstrated distinct genetic features compared to MCD-ABC-systemic DLBCL. (A) Boxplots comparing tumor mutation burden (TMB) of PCNSL compared to nodal ABC-systemic DLBCL and nodal ABC-systemic DLBCL classified as MCD by the LymphGen subgroup. PCNSL showed a significantly (P < 0.01, t-test) higher tumor mutation burden (TMB) relative to ABC-systemic DLBCL. However, when compared to just the MCD-ABC-systemic DLBCL genetic subtype, the TMB of PCNSL was similar. (B) Fisher’s exact tests were conducted between PCNSL (pink) and MCD-ABC-systemic DLBCL (green). The forest plot and corresponding bar plot revealed a significant enrichment (q < 0.1, Fisher’s exact test) in mutations in SOCS1, HNRNPU, GNAI2, GNA13, CXCR4, and CIITA in PCNSL compared to MCD-ABC-systemic DLBCL. (C) Copy number profiles highlighted amplifications and deletions per chromosome of PCNSL (top) and MCD-ABC-systemic DLBCL (bottom). (D-E) The figure represents a visualization of significant amplification and deletion peaks in the aggregate of PCNSL and MCD-ABC- systemic DLBCL, and genes associated with these peaks. The forest plot highlighted significantly enriched peaks in the aggregate of PCNSL and MCD-ABC-systemic DLBCL as defined by GISTIC v.2.0.23. Genes of interest associated with these peaks were annotated beside the peak.

As expected, due to the known enrichment of ABC types of large cell lymphomas in the MCD genetic group, in our comparison of sequencing data from ABC-PCNSL to ABC-systemic-DLBCL, we observed significant (q < 0.1, Fisher’s exact test) mutations in genes associated with the LymphGen MCD type including MYD88^L265P^, CD79B, and BTG1 in PCNSL (Supplemental Figure 8). In the strictest comparison, restricted to just MCD-ABC-systemic DLBCL, most differences between the groups were no longer observed (Supplemental Figure 9) except for increased mutations (q < 0.1, Fisher’s exact test) in SOCS1, HNRNPU, GNAI2, GNA13, CXCR4, and CIITA in PCNSL compared with MCD-ABC-systemic DLBCL (Figure 5B and Supplemental Figure 10). This shows a significant overlap in biology between MCD-ABC-systemic DLBCL and ABC-PCNSL.

Consistent with other studies that found increased JAK/STAT pathway usage in PCNSL, SOCS1 is a negative regulator of the JAK/STAT pathway ^29,30^. We also observed mutations affecting G-protein signaling (GNA13, GNAI2) and in CIITA, which encodes a regulator of class II major histocompatibility complex expression. Notably, aberrations in GNA13, GNAI2, and CIITA were associated with a mutational signature found in the mesenchymal-lymphoma microenvironment ^31^. Interestingly, we observed CXCR4 mutations in 14% of the PCNSL tumors versus 1% in the MCD-ABC-systemic DLBCL. CXCR4 mutations are rare in DLBCL, often associated with low-grade lymphoplasmacytic lymphoma, and have been implicated in increased resistance to ibrutinib ^32^. Finally, we identified unique mutations in the RNA-binding protein HNRNPU in 14% of the PCNSL, similar to the truncating mutations of HNRNPU previously identified in Burkitt lymphoma ^33^.

The copy number profiles of 89 MCD-ABC-systemic DLBCL were compared to 16 PCNSL tumors with higher purity using GISTIC v.2.0.23 to determine recurrent CNA. The overall profile of PCNSL resembled that of MCD-ABC-systemic DLBCL, with some key differences notably, unique arm-level deletions affecting chromosome 10p in PCNSL overall (q < 0.1, Fisher’s exact test) (Figure 5C). In contrast, unique arm-level amplifications were observed at chromosome 3q in the MCD-ABC-systemic DLBCL. Chromosome 6q deletions frequently occurred in both groups, although 6q21 deletions containing PRDM1 occurred at a higher frequency in PCNSL (88%) compared to MCD-ABC-systemic DLBCL (47%) (Figure 5D and E). Similarly, we observed a higher incidence of focal deletions at 19p13.2 containing MEF2B (38%) and at 3p21.31 containing RHOA (38%) in PCNSL compared to MCD-ABC-systemic DLBCL (11% and 6%, respectively). Focal amplifications at 14q24.1 (ZFP36L1), 11q13.1 (MALAT1), and 1p31.3 (PDE4B), were observed at a higher prevalence in PCNSL compared to MCD-ABC-systemic DLBCL. Other copy number alterations commonly observed in MCD-ABC-systemic DLBCL including deletions in 12p13.2 (ETV6), 1p13.1 (CD58), and 1p36.31 (NOL9, KLHL21), were detected at similar levels in PCNSL (Figure 5E). After manual curation the CDKN2A locus, we found similar levels of CDKN2A deletions in MCD-ABC-systemic DLBCL 43% (38/89) compared to PCNSL 56% (9/16).

PCNSL cell lines show variable sensitivity to chemotherapy, BTK, FGFR, and PI3K inhibitors

To evaluate potential associations between GEP pathway score and drug response, two PCNSL cell lines (TK and HKBML) and three systemic DLBCL cell lines (TMD8, OCI-Ly3, and HBL-1), all verified as ABC subtypes by Lymph2Cx, were run using the PanCancer Pathways panel (Figure 6 A, duplicates for each cell line). Three of the five cell lines in the experimental panel, HKBML, TMD8, and HBL-1, showed higher pathway scores across most of the cancer-related pathways than the TK and OCI-LY3 lines (Figure 6A).

**Figure 6.**
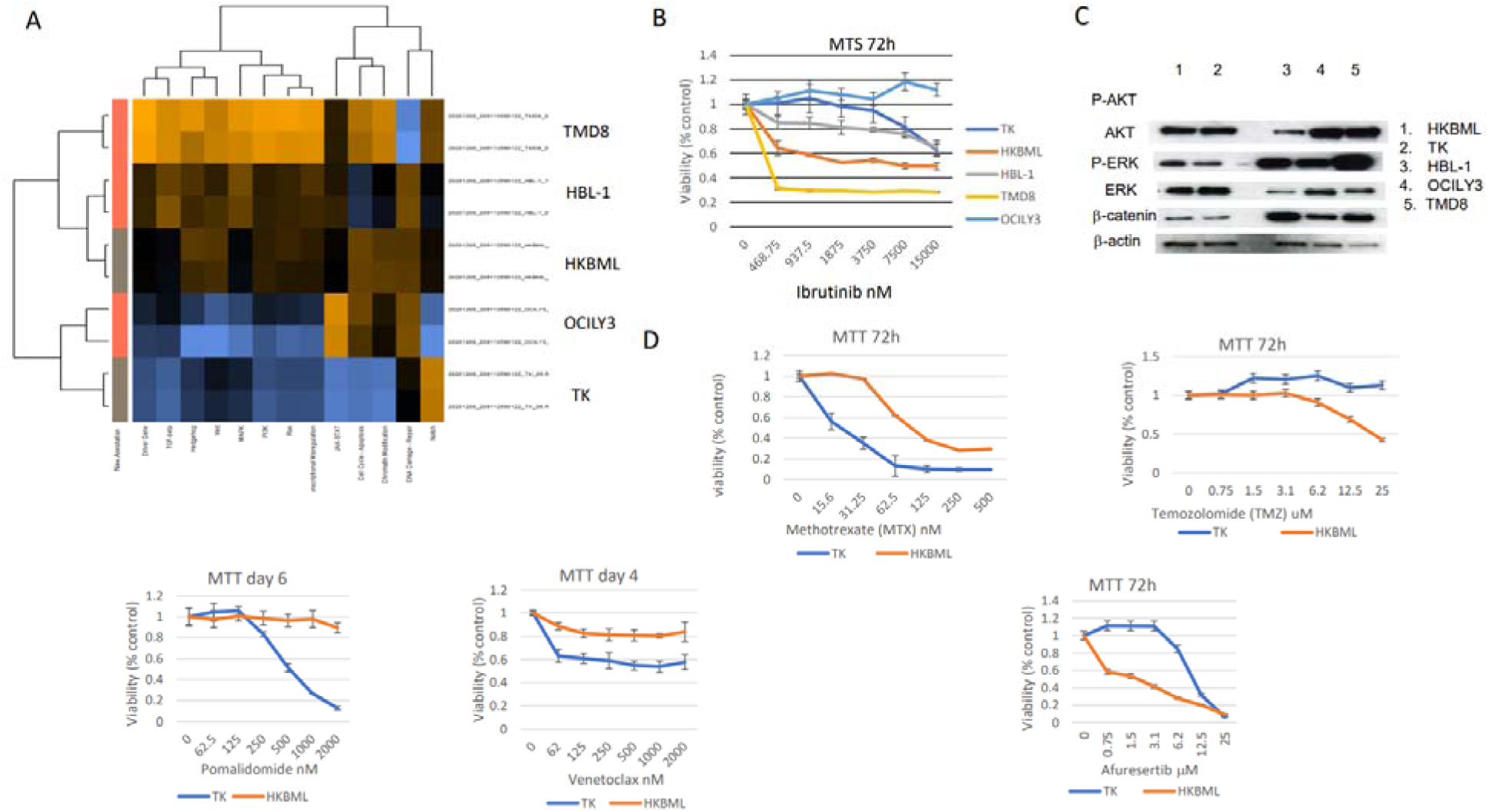
Two PCNSL cell lines had variable sensitivity to conventional anti-lymphoma chemotherapy, BTK, FGFR, and PI3K inhibitors. (A) Two ABC-PCNSL cell lines (TK and HKBML) and three ABC-systemic DLBCL cell lines were tested on a PanCancer Pathways panel. A heatmap of “pathway scores” indicated some similarity between the PCNSL cell lines and the two transcriptional subgroups (P1 and P2) identified in PCNSL patient samples. (B) MTS assays were performed 72 hours after five DLBCL cell lines were treated with a specific BTK inhibitor, Ibrutinib. The results of a representative experiment in triplicate are shown. (C) Western blotting reveals potential correlation between activated AKT and Ibrutinib sensitivities in the tested cell lines. (D) The PCNSL cell lines were further tested by MTT assay for responses to four clinical drugs for PCNSL and lymphoma (MTX, TMZ, pomalidomide and venetoclax) and to two inhibitors against AKT (Afuresertib) or FGFR kinase (BGJ398). (E) Combination of MTX with FGF inhibitor generated a potentially synergistic antitumor activity on HKBML.

Next, we tested drug responses of all 5 lines to ibrutinib, which targets BTK and has been used to treat ABC-systemic DLBCL patients ^34^. As expected, TK and OCI-Ly3 both showed resistance to ibrutinib (Figure 6B). Interestingly, TK and OCI-LY3 both showed lower GEP “pathway scores” than other cell lines. Consistent with the “pathway score” analysis, western blotting of the two PCNSL cell lines demonstrated HKBML had more activated PI3K and ERK signaling than TK (Figure 6C). Further testing with additional drugs/compounds on those two PCNSL cell lines demonstrated differential results. Although this is a limited study (due to lack of PCNSL lines in the published literature), our results showed that, compared to the TK cell line, HKBML (with a higher pathway score) was less responsive to methotrexate (inhibits purine and pyrimidine synthesis, commonly used to treat PCNSL), pomalidomide (as an example immunomodulatory drug), venetoclax (anti-BCL2 mechanism), and an FGF inhibitor (BGJ 398), was more sensitive to TMZ (alkylating agent), and afuresertib (PI3 kinase pathway inhibitor) (Figure 6D). Overall, these findings suggest that the discovered transcriptional heterogeneity of PCNSL may impact responsiveness to therapy.

## Discussion

Evaluating gene expression profiles for CNS lymphoma is more challenging than for systemic DLBCL due to small sample sizes, variable cell density, variable infiltration by immune effector cells, and background brain tissue ^35^. We minimized this by macro-dissecting samples to reach a minimum of 60% tumor. An additional feature of this work is the focus on cases that were both EBV(-) and HIV(-)in order to create a rigorous, uniform cohort. Secondary lymphomas involving the CNS were not a part of the current study group but could be the focus of future studies. The main limitations of this project include the acknowledged heterogeneity in tumor purity (which we attempted to address with macro-dissection and in various analyses such as CNA where possible), small cohort size (yet relatively large for this rare and hard to biopsy disease) and therefore limited ability to address multiple testing and lack of replication. After an extensive search, only 2 acknowledged PCNSL lines were found in the literature and cell line catalogs, limiting the in vitro work.

Previously, Tun et al used GEP to compare PCNSL to other types of lymphoma and found a unique CNS signature in which there was significant differential expression of multiple extracellular matrix and adhesion-related pathways ^36^. Specifically, the secreted phosphoprotein 1 (SPP1) gene was highly upregulated, while the nicotinamide N-methyltransferase (NNMT) and vascular endothelial growth factor (VEGF) genes were downregulated. SPP1 reportedly has a role in osteoclast attachment to bone matrix, but also as a cytokine that upregulates expression of interferon-gamma and interleukin-12, the latter involved in both T and NK cell activation. Similar findings were also reported by Rubinstein et al in their microarray study ^35^. Another GEP study found that PCNSL did not differ markedly from systemic DLBCL, but did show that SPP1 is highly expressed in PCNSL B cells ^23^. We had similar findings and confirmed over-expression of SPP1 and down-regulation of VEGFC in PCNSL (Figure 1C and Supplemental data 2). The current study additionally identified new genes and pathways that were differentially expressed between ABC-PCNSL and ABC-systemic DLBCL samples, which may help in identifying new biology and therapeutic strategies. For example, we found that MCL-1 expression is higher and BCL-2 is lower in ABC-PCNSL samples compared with ABC-systemic DLBCL, giving rise to the speculation that ABC-PCNSL patients may be less responsive to Venetoclax and inhibition of MCL-1 could be more effective.

We observed unique mutations in PCNSL including GNA13, GNAI2, and CIITA, likely representing a lymphoma tumor microenvironment mutational signature ^31^. We also observed CXCR4 mutations in 14% of PCNSL; unlike in lymphoplasmacytic lymphoma in which 43% of CXCR4 mutations were S338X mutations, the majority of mutations in PCNSL occurred in the transmembrane domain of CXCR4 with consequent implications for ibrutinib (commonly used to treat ABC types of DLBCL) resistance ^37^. In MCD-ABC-PCNSL, mutations in SOCS1, a negative regulator of the JAK-STAT pathway, were also enriched.

Chapuy et al aimed to identify genetic features of PCNSL and primary testicular lymphoma using WES. They identified several recurrent mutations involved in NF-kB signaling pathway, including MYD88, CD79B, and PIM1, identical to our own findings. Functional studies confirmed the role of MYD88 mutations in activating NF-kB pathway and promoting survival of PCNSL cells ^38^. Another study using whole genome and RNA sequencing to characterize the genetic landscape of both primary and secondary large B-cell lymphomas, EBV(+) and EBV(-), in the CNS found significantly more single nucleotide variants (SNVs) and indels compared to systemic DLBCL, even in the intronic and intergenic regions ^39^. In addition, that group of investigators found recurrent mutations in JAK-STAT, NF-kB, and B-cell receptor signaling pathways, alongside other mutations also found in other ABC-systemic DLBCL such as MYD88, CD79B, as well as deletions of MTAP and CDKN2A at 9p21 and CDKN2D at 19p13 in the PCNSL cases. PCNSL also had more frequent deletion of the human leukocyte antigen (HLA) type I D locus (HLA-D) at 6p21, implicating immune evasion in lymphomagenesis. In fact, homozygous deletions of the HLA type I and II loci have long been known to be more common in PCNSL compared to systemic DLBCL ^40^. Other features of PCNSL included mutational signatures related to DNA replication and mitosis. In addition, TERT gene expression (but not telomere content) and IgM expression (as compared to IgG expression) were both significantly higher in PCNSL compared to the secondary cases. Finally, the EBV(+) cases (primary and secondary) reportedly lacked recurrent mutational hotspots apart from immunoglobulin and HLA-DRB loci ^39^.

Due to small sample sizes and limited amounts of RNA extracted from PCNSL samples, we could not perform duplicate runs of patient samples or perform further validation using RT-PCR or immunostaining. However, previous studies from our group and others have repeatedly demonstrated that the data generated from FFPE samples using nCounter technology correlate with that generated from matched fresh frozen material or QPCR data ^10,41,42^. Analysis of a limited panel of two PCNSL cell lines with different GEP “pathway scores” correlated with their responses to different drugs and inhibitors and generally correlated with activation of certain pathways detected by GEP, such as PI3 signaling. PI3K inhibitors have been evaluated in PCNSL. The first trial was of buparlisib, which closed prematurely due to limited response ^43^. Another study evaluated PQR309, a novel dual PI3K/mTOR inhibitor, but the results did not seem promising, and data are yet published (NCT02669511, NCT03120000). An attractive approach is combining therapies targeting different pathways. Copanlisib, a pan-PI3K inhibitor is currently being evaluated in combination with ibrutinib in relapsed PCNSL in a phase Ib/II trial (NCT03581942).

Altogether, these findings add to the existing genomic and transcriptional landscape of PCNSL and suggest heterogeneity within the disease with possible targetable biology.

## Data Availability

All data produced in the present study are available upon reasonable request to the authors.

## Acknowledgement

The authors wish to acknowledge the gene expression data shared by Dr. Manuel Montesinos- Rongen as well as the Lymphoma & Leukemia Molecular Profiling Project for their critical review of the data.

## References

1. Hilal T, Maguire A, Kosiorek HE, Rimsza LM, Rosenthal AC. Clinical features and cell of origin subtyping using gene expression profiling in HIV-negative patients with primary central nervous system lymphoma. Leuk Lymphoma. 2019;60(14):3581–3583. doi:10.1080/10428194.2019.1633637

2. WHO Classification of Tumours Editorial Board. Haematolymphoid Tumours. Vol 11. 5th ed. International Agency for Research on Cancer; 2024. https://publications.iarc.who.int/637

3. Swerdlow SH, Campo E, Pileri SA, et al. The 2016 revision of the World Health Organization classification of lymphoid neoplasms. Blood. 2016;127(20):2375-2390. doi:10.1182/blood-2016-01-643569

4. Mendez JS, Ostrom QT, Gittleman H, et al. The elderly left behind—changes in survival trends of primary central nervous system lymphoma over the past 4 decades. Neuro-Oncol. 2018;20(5):687–694. doi:10.1093/neuonc/nox187

5. Hilal T. Primary central nervous system lymphoma: Consensus, controversies, and future directions. Adv CELL GENE Ther. 2020;3(3):e82. doi:10.1002/acg2.82

6. Houillier C, Soussain C, Ghesquières H, et al. Management and outcome of primary CNS lymphoma in the modern era. Neurology. 2020;94(10):e1027–e1039. doi:10.1212/WNL.0000000000008900

7. Cerhan JR, Link BK, Habermann TM, et al. Cohort Profile: The Lymphoma Specialized Program of Research Excellence (SPORE) Molecular Epidemiology Resource (MER) Cohort Study. Int J Epidemiol. 2017;46(6):1753–1754i. doi:10.1093/ije/dyx119

8. Fujimoto K, Shinojima N, Hayashi M, Nakano T, Ichimura K, Mukasa A. Histone deacetylase inhibition enhances the therapeutic effects of methotrexate on primary central nervous system lymphoma. Neuro-Oncol Adv. 2020;2(1):vdaa084. doi:10.1093/noajnl/vdaa084

9. Miyoshi I, Kubonishi I, Yoshimoto S, et al. Characteristics of a brain lymphoma cell line derived from primary intracranial lymphoma. Cancer. 1982;49(3):456–459. doi:10.1002/1097-0142(19820201)49:3<456::AID-CNCR2820490311>3.0.CO;2-K

10. Scott DW, Wright GW, Williams PM, et al. Determining cell-of-origin subtypes of diffuse large B-cell lymphoma using gene expression in formalin-fixed paraffin-embedded tissue. Blood. 2014;123(8):1214–1217. doi:10.1182/blood-2013-11-536433

11. Li H, Durbin R. Fast and accurate short read alignment with Burrows–Wheeler transform. Bioinformatics. 2009;25(14):1754–1760. doi:10.1093/bioinformatics/btp324

12. Kim S, Scheffler K, Halpern AL, et al. Strelka2: fast and accurate calling of germline and somatic variants. Nat Methods. 2018;15(8):591–594. doi:10.1038/s41592-018-0051-x

13. McKenna A, Hanna M, Banks E, et al. The Genome Analysis Toolkit: A MapReduce framework for analyzing next-generation DNA sequencing data. Genome Res. 2010;20(9):1297–1303. doi:10.1101/gr.107524.110

14. Wilm A, Aw PPK, Bertrand D, et al. LoFreq: a sequence-quality aware, ultra-sensitive variant caller for uncovering cell-population heterogeneity from high-throughput sequencing datasets. Nucleic Acids Res. 2012;40(22):11189-11201. doi:10.1093/nar/gks918

15. Wright GW, Huang DW, Phelan JD, et al. A Probabilistic Classification Tool for Genetic Subtypes of Diffuse Large B Cell Lymphoma with Therapeutic Implications. Cancer Cell. 2020;37(4):551–568.e14. doi:10.1016/j.ccell.2020.03.015

16. Riester M, Singh AP, Brannon AR, et al. PureCN: copy number calling and SNV classification using targeted short read sequencing. Source Code Biol Med. 2016;11(1):13. doi:10.1186/s13029-016-0060-z

17. Mermel CH, Schumacher SE, Hill B, Meyerson ML, Beroukhim R, Getz G. GISTIC2.0 facilitates sensitive and confident localization of the targets of focal somatic copy-number alteration in human cancers. Genome Biol. 2011;12(4):R41. doi:10.1186/gb-2011-12-4-r41

18. Talevich E, Shain AH, Botton T, Bastian BC. CNVkit: Genome-Wide Copy Number Detection and Visualization from Targeted DNA Sequencing. PLOS Comput Biol. 2016;12(4):e1004873. doi:10.1371/journal.pcbi.1004873

19. Jia W, Li H, Li S, Chen L, Li SC. Oviz-Bio: a web-based platform for interactive cancer genomics data visualization. Nucleic Acids Res. 2020;48(W1):W415–W426. doi:10.1093/nar/gkaa371

20. Chapuy B, Stewart C, Dunford AJ, et al. Molecular subtypes of diffuse large B cell lymphoma are associated with distinct pathogenic mechanisms and outcomes. Nat Med. 2018;24(5):679–690. doi:10.1038/s41591-018-0016-8

21. Reddy A, Zhang J, Davis NS, et al. Genetic and Functional Drivers of Diffuse Large B Cell Lymphoma. Cell. 2017;171(2):481–494.e15. doi:10.1016/j.cell.2017.09.027

22. Schmitz R, Wright GW, Huang DW, et al. Genetics and Pathogenesis of Diffuse Large B-Cell Lymphoma. N Engl J Med. 2018;378(15):1396–1407. doi:10.1056/NEJMoa1801445

23. Montesinos-Rongen M, Brunn A, Bentink S, et al. Gene expression profiling suggests primary central nervous system lymphomas to be derived from a late germinal center B cell. Leukemia. 2008;22(2):400–405. doi:10.1038/sj.leu.2405019

24. Zhu Q, Wang J, Zhang W, et al. Whole-Genome/Exome Sequencing Uncovers Mutations and Copy Number Variations in Primary Diffuse Large B-Cell Lymphoma of the Central Nervous System. Front Genet. 2022;13. doi:10.3389/fgene.2022.878618

25. Arthur SE, Jiang A, Grande BM, et al. Genome-wide discovery of somatic regulatory variants in diffuse large B-cell lymphoma. Nat Commun. 2018;9(1):4001. doi:10.1038/s41467-018-06354-3

26. Dreval K, Hilton LK, Cruz M, et al. Genetic subdivisions of follicular lymphoma defined by distinct coding and noncoding mutation patterns. Blood. 2023;142(6):561–573. doi:10.1182/blood.2022018719

27. Runge HFP, Lacy S, Barrans S, et al. Application of the LymphGen classification tool to 928 clinically and genetically-characterised cases of diffuse large B cell lymphoma (DLBCL). Br J Haematol. 2021;192(1):216–220. doi:10.1111/bjh.17132

28. Caeser R, Walker I, Gao J, et al. Acquired CARD11 Mutation Promotes BCR Independence in Diffuse Large B Cell Lymphoma. JCO Precis Oncol. 2021;(5):145–152. doi:10.1200/PO.20.00360

29. Liau NPD, Laktyushin A, Lucet IS, et al. The molecular basis of JAK/STAT inhibition by SOCS1. Nat Commun. 2018;9(1):1558. doi:10.1038/s41467-018-04013-1

30. Mondello P, Cuzzocrea S, Arrigo C, Pitini V, Mian M, Bertoni F. STAT6 activation correlates with cerebrospinal fluid IL-4 and IL-10 and poor prognosis in primary central nervous system lymphoma. Hematol Oncol. 2020;38(1):106–110. doi:10.1002/hon.2679

31. Kotlov N, Bagaev A, Revuelta MV, et al. Clinical and Biological Subtypes of B-cell Lymphoma Revealed by Microenvironmental Signatures. Cancer Discov. 2021;11(6):1468–1489. doi:10.1158/2159-8290.CD-20-0839

32. Castillo JJ, Xu L, Gustine JN, et al. CXCR4 mutation subtypes impact response and survival outcomes in patients with Waldenström macroglobulinaemia treated with ibrutinib. Br J Haematol. 2019;187(3):356–363. doi:10.1111/bjh.16088

33. Thomas N, Dreval K, Gerhard DS, et al. Genetic subgroups inform on pathobiology in adult and pediatric Burkitt lymphoma. Blood. 2023;141(8):904–916. doi:10.1182/blood.2022016534

34. Grommes C, Pastore A, Palaskas N, et al. Ibrutinib Unmasks Critical Role of Bruton Tyrosine Kinase in Primary CNS Lymphoma. Cancer Discov. 2017;7(9):1018–1029. doi:10.1158/2159-8290.CD-17-0613

35. Rubenstein JL, Fridlyand J, Shen A, et al. Gene expression and angiotropism in primary CNS lymphoma. Blood. 2006;107(9):3716–3723. doi:10.1182/blood-2005-03-0897

36. Tun HW, Personett D, Baskerville KA, et al. Pathway analysis of primary central nervous system lymphoma. Blood. 2008;111(6):3200–3210. doi:10.1182/blood-2007-10-119099

37. Gustine JN, Xu L, Tsakmaklis N, et al. CXCR4S338X clonality is an important determinant of ibrutinib outcomes in patients with Waldenström macroglobulinemia. Blood Adv. 2019;3(19):2800–2803. doi:10.1182/bloodadvances.2019000635

38. Chapuy B, Roemer MGM, Stewart C, et al. Targetable genetic features of primary testicular and primary central nervous system lymphomas. Blood. 2016;127(7):869–881. doi:10.1182/blood-2015-10-673236

39. Radke J, Ishaque N, Koll R, et al. The genomic and transcriptional landscape of primary central nervous system lymphoma. Nat Commun. 2022;13(1):2558. doi:10.1038/s41467-022-30050-y

40. Riemersma SA, Jordanova ES, Schop RFJ, et al. Extensive genetic alterations of the HLA region, including homozygous deletions of HLA class II genes in B-cell lymphomas arising in immune-privileged sites. Blood. 2000;96(10):3569–3577. doi:10.1182/blood.V96.10.3569

41. Chen X, Deane NG, Lewis KB, et al. Comparison of Nanostring nCounter® Data on FFPE Colon Cancer Samples and Affymetrix Microarray Data on Matched Frozen Tissues. PLOS ONE. 2016;11(5):e0153784. doi:10.1371/journal.pone.0153784

42. Ryan MT, Martinez C, Jahns H, et al. The comparative performance of a custom Canine NanoString® panel on FFPE and snap frozen liver biopsies. Res Vet Sci. 2023;159:225–231. doi:10.1016/j.rvsc.2023.04.023

43. Grommes C, Pentsova E, Nolan C, Wolfe J, Mellinghoff IK, Deangelis L. 335P - Phase II study of single agent buparlisib in recurrent/refractory primary (PCNSL) and secondary CNS lymphoma (SCNSL). Ann Oncol. 2016;27:vi106. doi:10.1093/annonc/mdw367.13

