## supplemental figures 1-10 for "Primary Central Nervous System Lymphoma Tumor Biopsies Show Heterogeneity in Gene Expression Profiles and Genetic Subtypes"

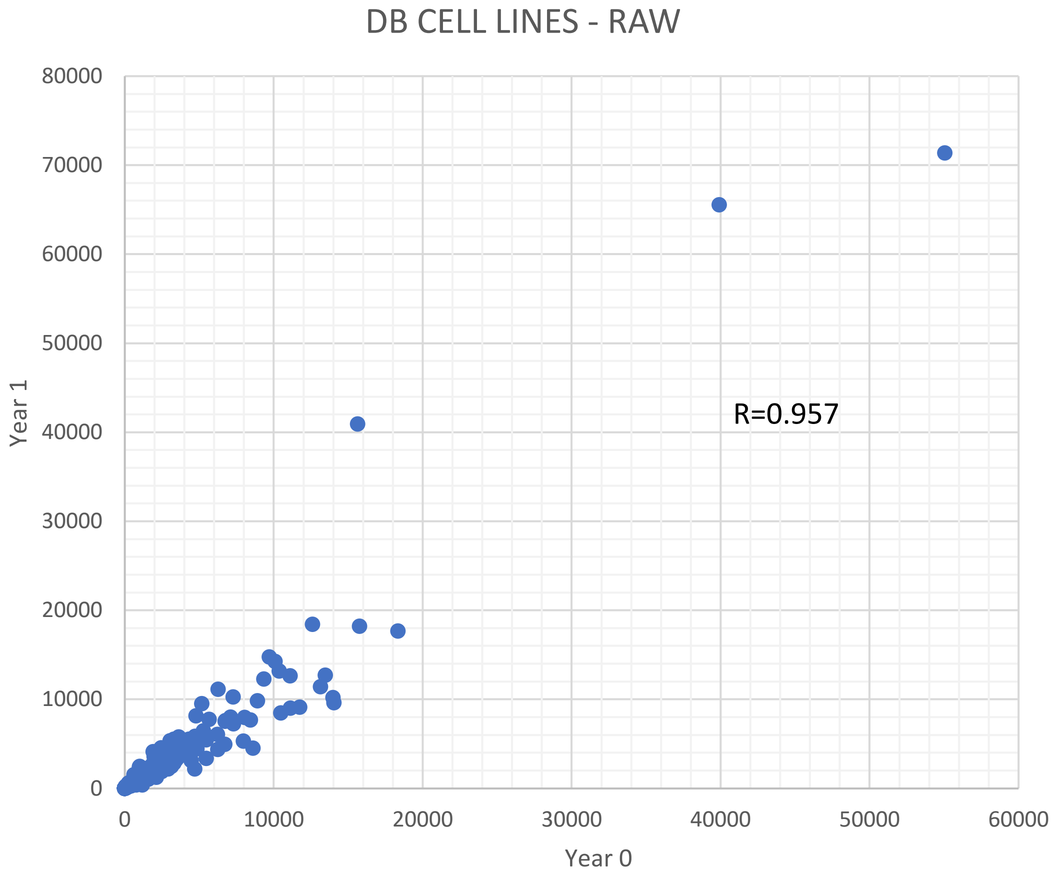


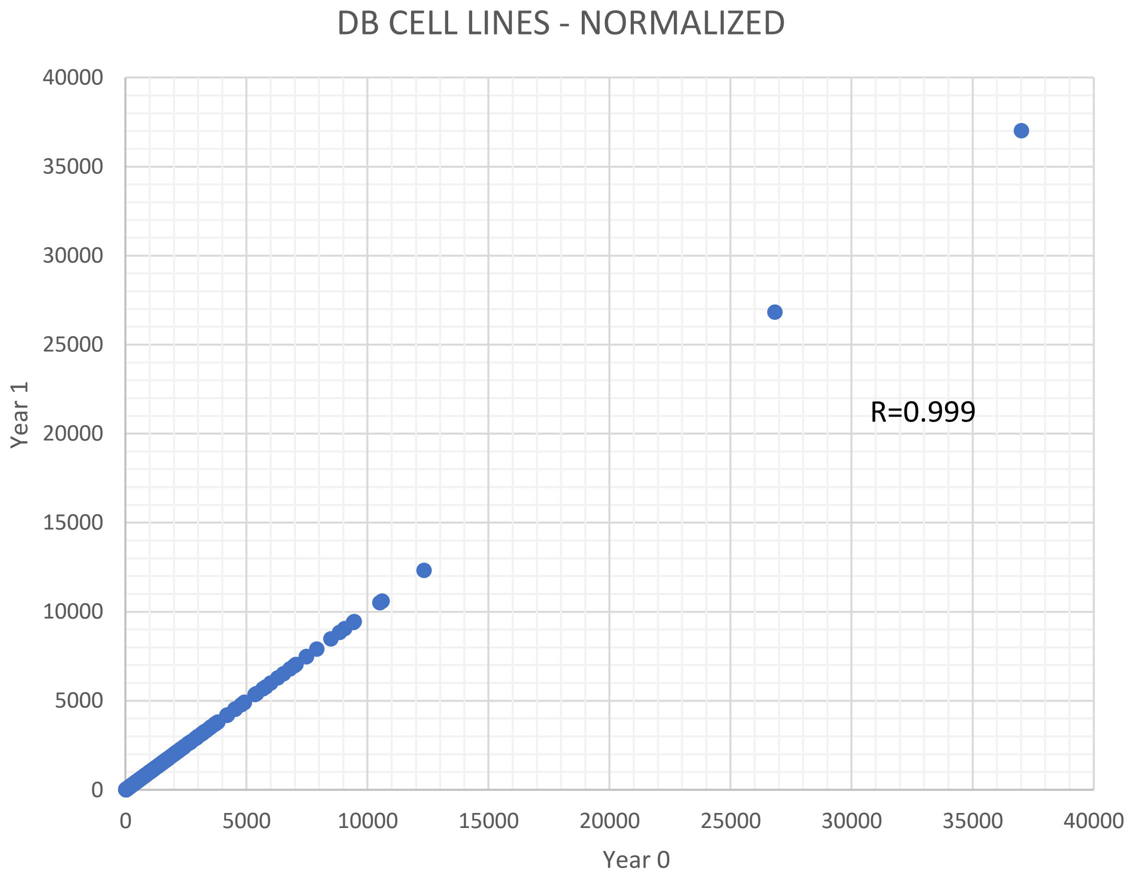


**Supplemental figure 1**

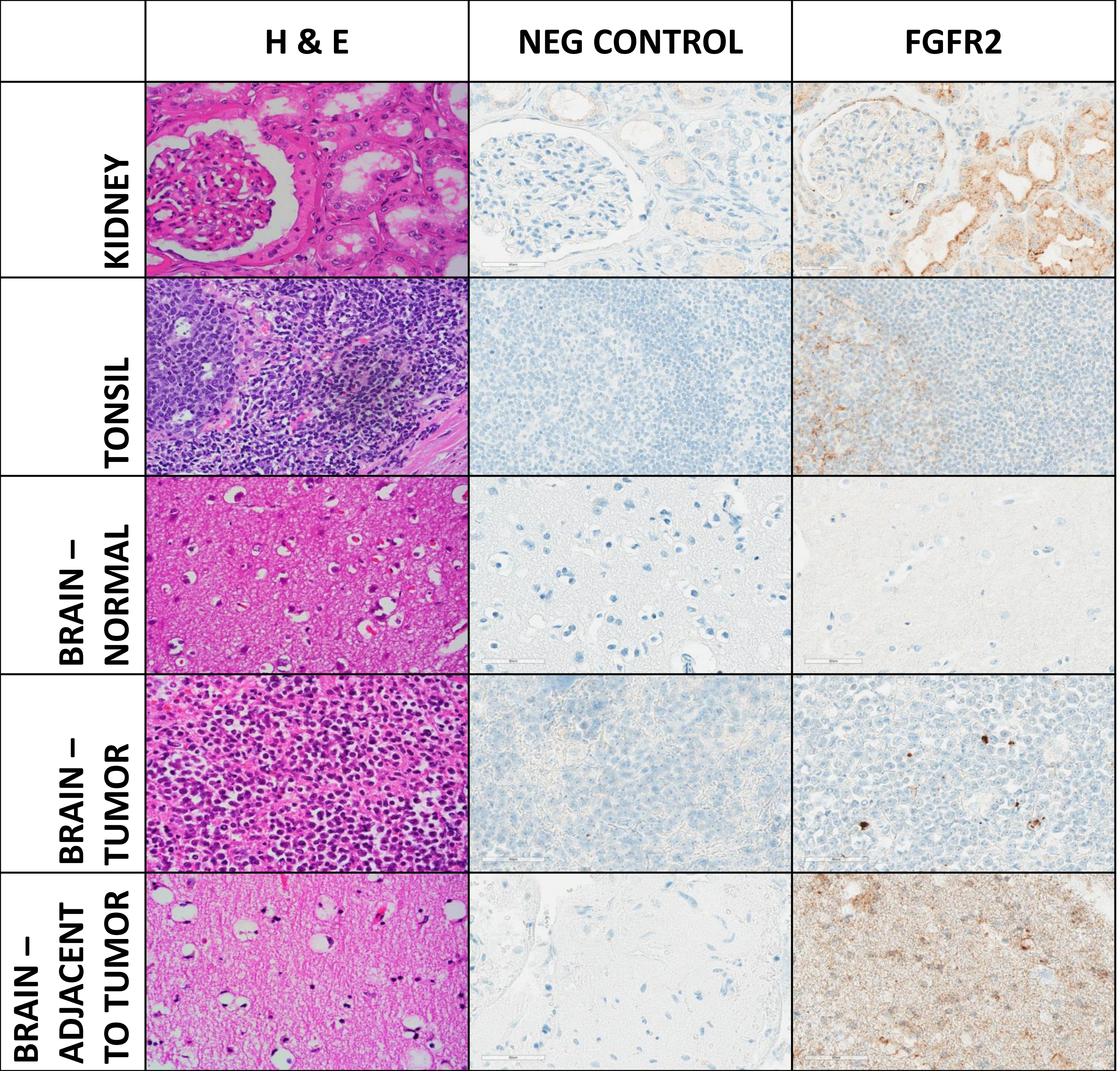


**Supplemental figure 2**

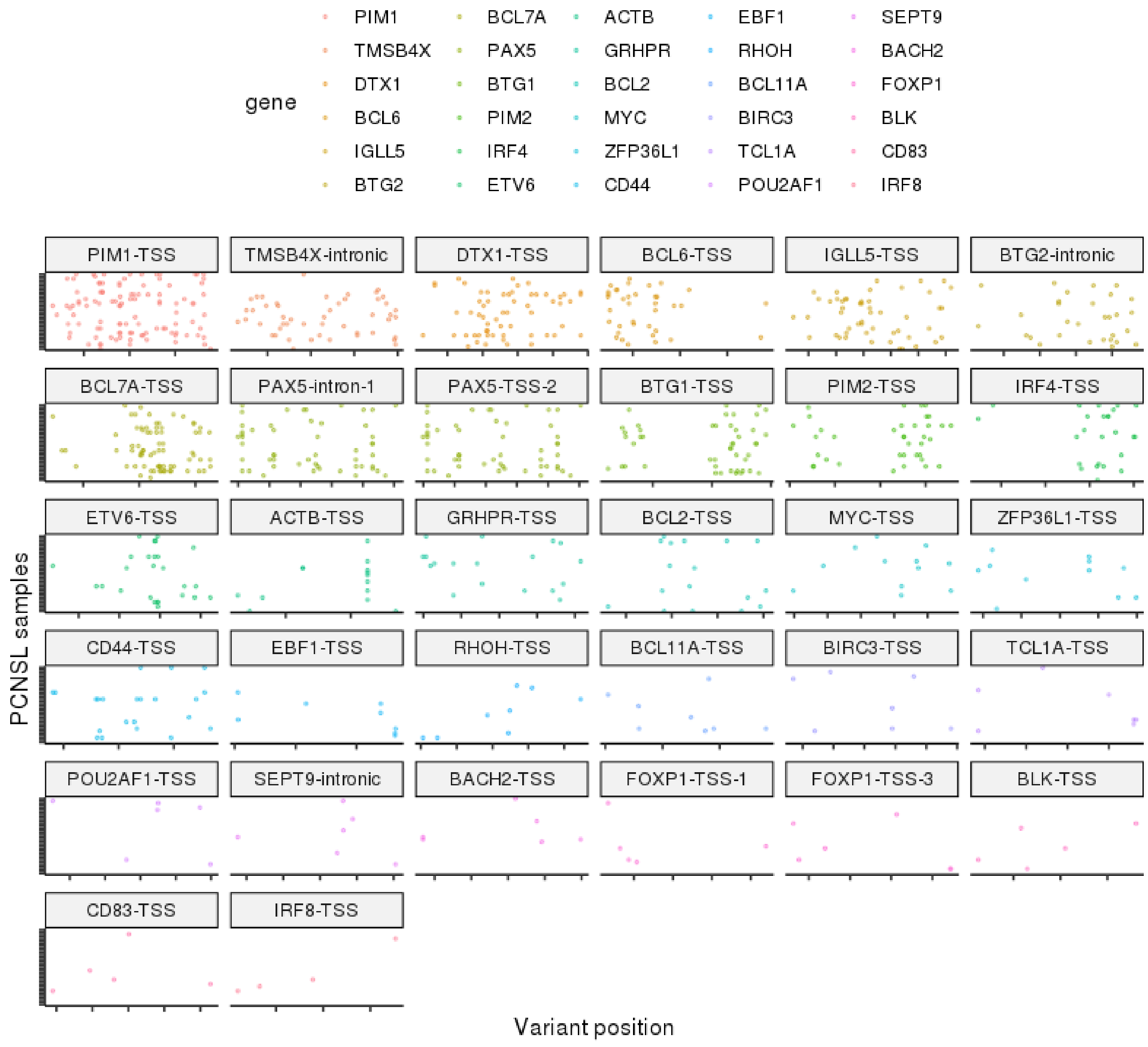


**Supplemental figure 3**

No differences between transcriptional subgroups

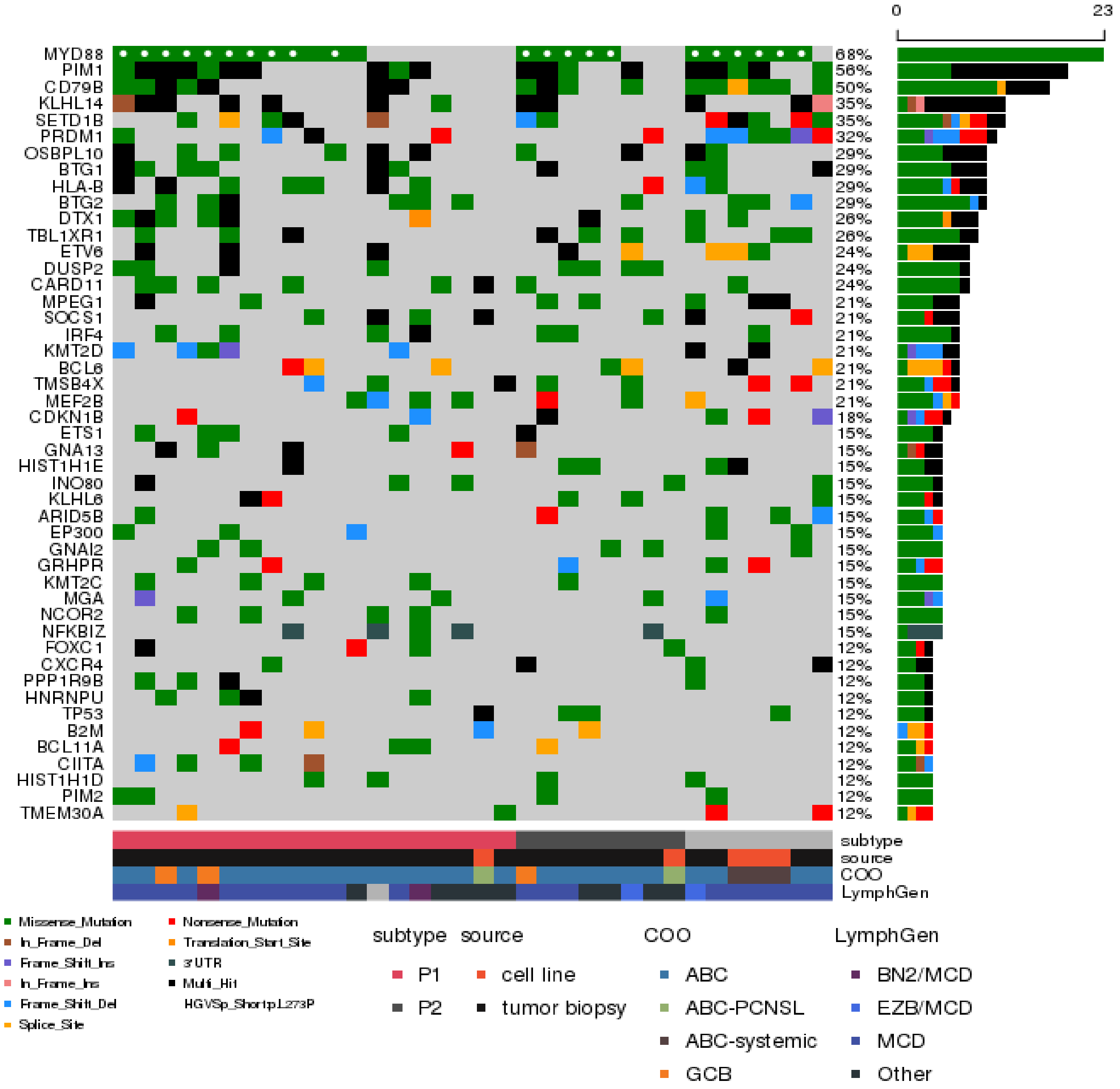


| NanoString ID | Global ID (SPEC #) | SEX | MER ID | Lymph2Cx Assays | LymphGen | suptype |
|---|---|---|---|---|---|---|
| P34 | 5447 | M | IA6787 | ABC | BN2 | P1 |
| P35 | 5448 | M | IA6790 | GCB | BN2 | P1 |
| P2 | 5415 | M | IA6618 | ABC | MCD | P1 |
| P3 | 5416 | M | IA6339 | ABC | MCD | P1 |
| P4 | 5417 | F | MC4654 | ABC | MCD | P1 |
| P7 | 5420 | F | IA6364 | ABC | MCD | P1 |
| P10 | 5423 | M | IA1685 | ABC | MCD | P1 |
| P17 | 5430 | M | MC3175 | ABC | MCD | P1 |
| P27 | 5440 | F | MC7549 | GCB | MCD | P1 |
| P30 | 5443 | M | MC7668 | ABC | MCD | P1 |
| P36 | 5449 | F | IA6735 | ABC | MCD | P1 |
| P1 | 5414 | F | MC5939 | ABC | MCD/ST2 | P1 |
| P8 | 5421 | F | IA828 | ABC | Other | P1 |
| P11 | 5424 | M | IA2333 | ABC | Other | P1 |
| P15 | 5428 | M | IA3037 | ABC | Other | P1 |
| P19 | 5432 | F | MC2802 | ABC | Other | P1 |
| P23 | 5436 | M | MC5667 | ABC | Other | P1 |
| P29 | 5442 | M | MC7419 | ABC | Other | P1 |
| P14 | 5427 | M | IA2871 | ABC | | P1 |
| P26 | 5439 | M | MC7215 | ABC | EZB/MCD | P2 |
| P9 | 5422 | M | IA1457 | ABC | MCD | P2 |
| P31 | 5444 | F | MC3963 | GCB | MCD | P2 |
| P37 | 5450 | F | IA7348 | ABC | MCD | P2 |
| P5 | 5418 | F | IA5861 | ABC | Other | P2 |
| P32 | 5445 | F | MC4070 | ABC | Other | P2 |
| P33 | 5446 | F | MC2327 | ABC | Other | P2 |
| P20 | 5433 | F | MC392 | ABC | | P2 |
| P24 | 5437 | M | MC3474 | ABC | | P2 |
| P25 | 5438 | M | MC3916 | ABC | EZB/MCD | |
| P16 | 5429 | M | IA5731 | ABC | MCD | |
| P28 | 5441 | F | MC6892 | ABC | MCD | |

**Supplemental figure 4**

Dotted line represents
cut-off (at 0.0002%)

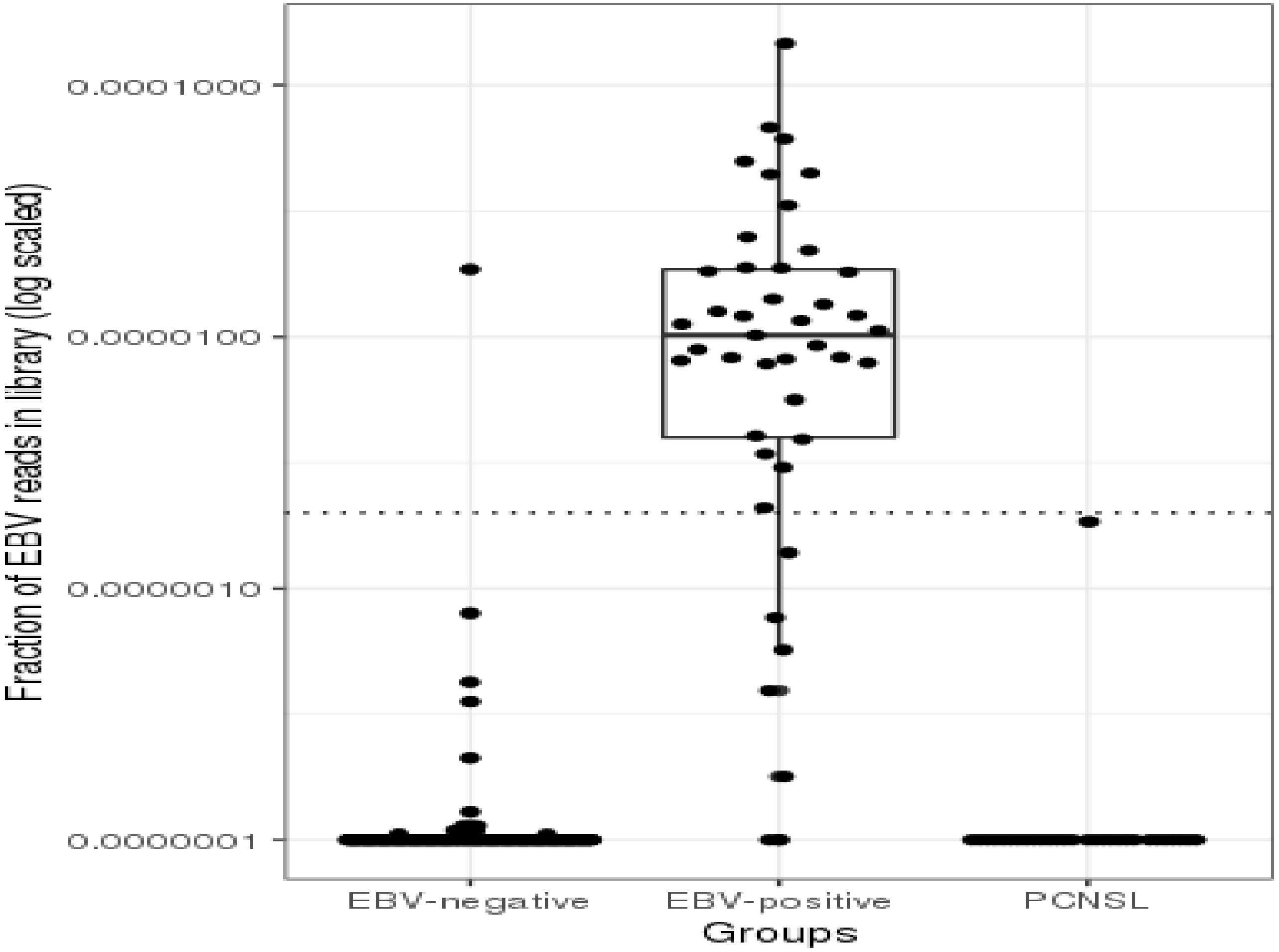


**Supplemental figure 5**

GISTIC of PCNSL samples with purity > 0.4 (n=16)

q < 0.1

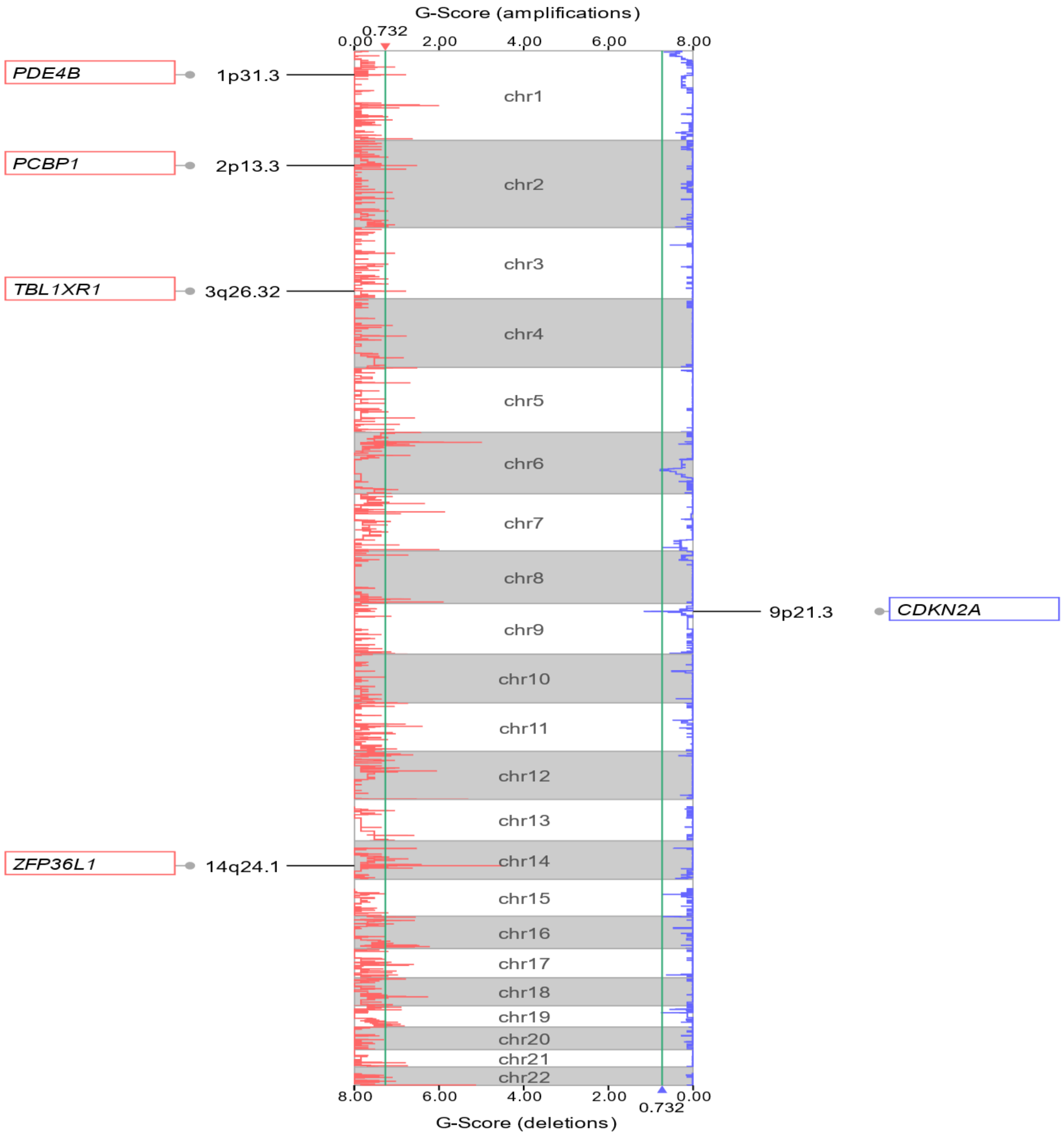


**Supplemental figure 6**

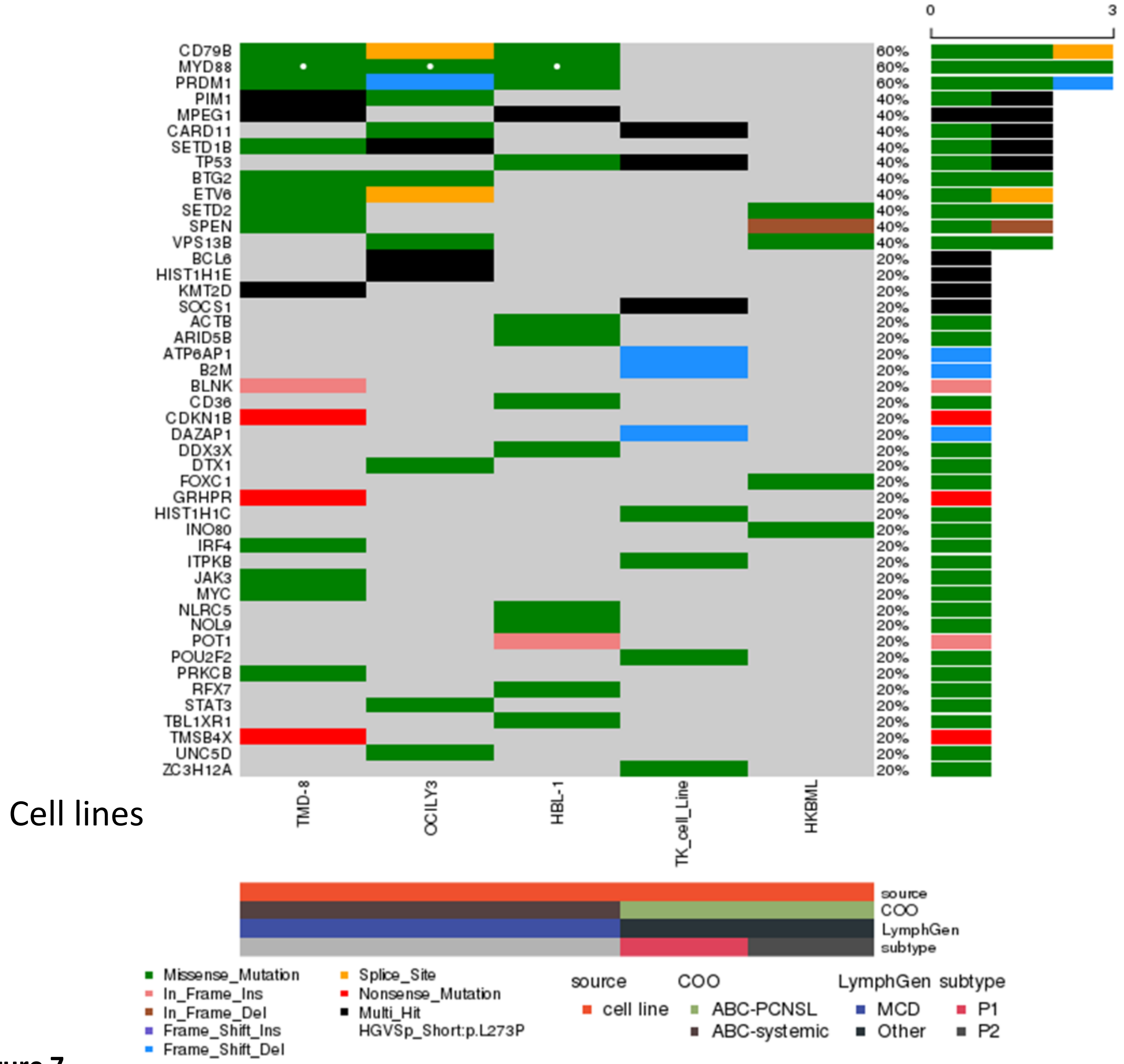


**Supplemental figure 7**

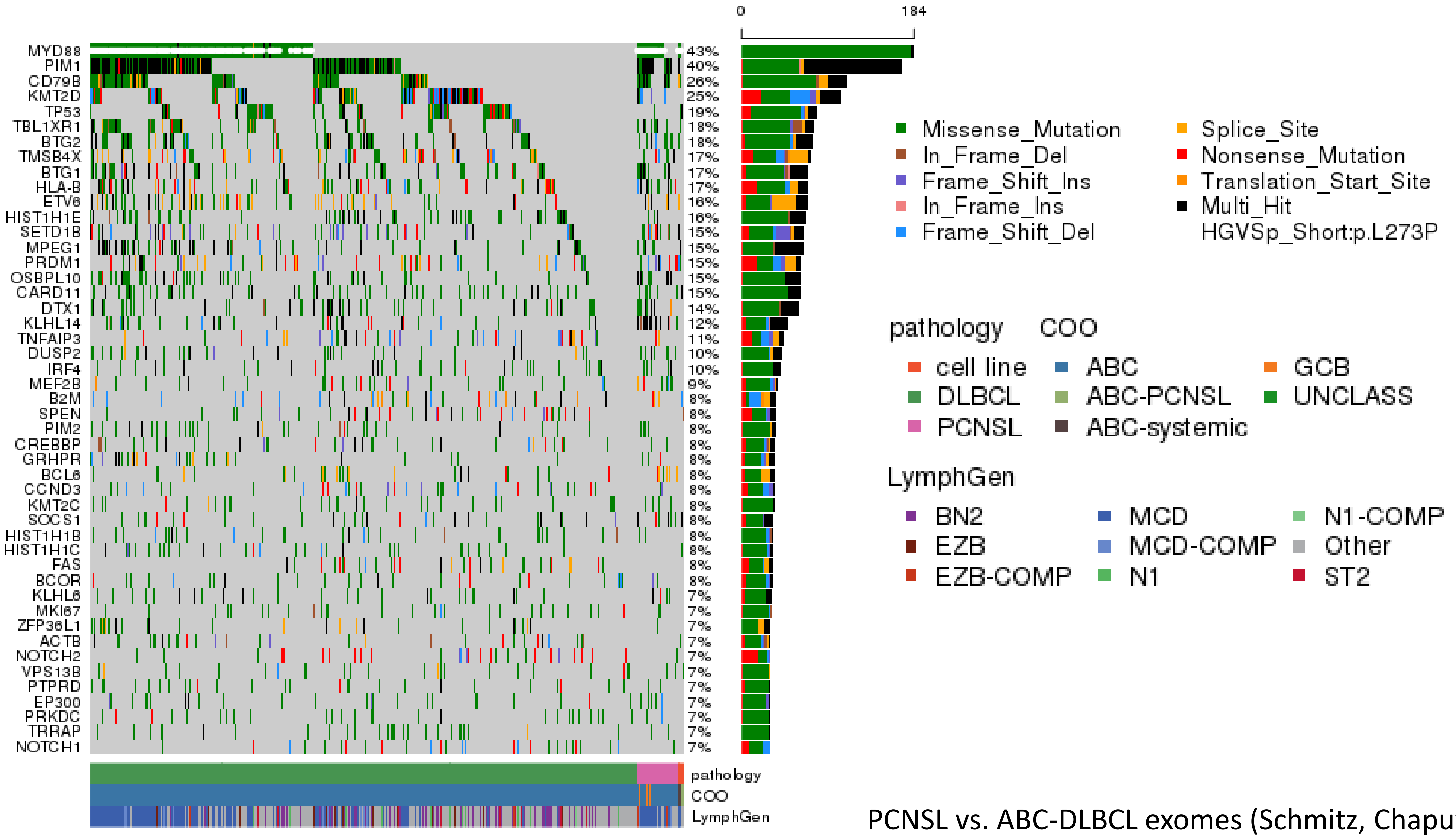


PCNSL vs. ABC-DLBCL exomes (Schmitz, Chapuy, Reddy)

**Supplemental figure 8**

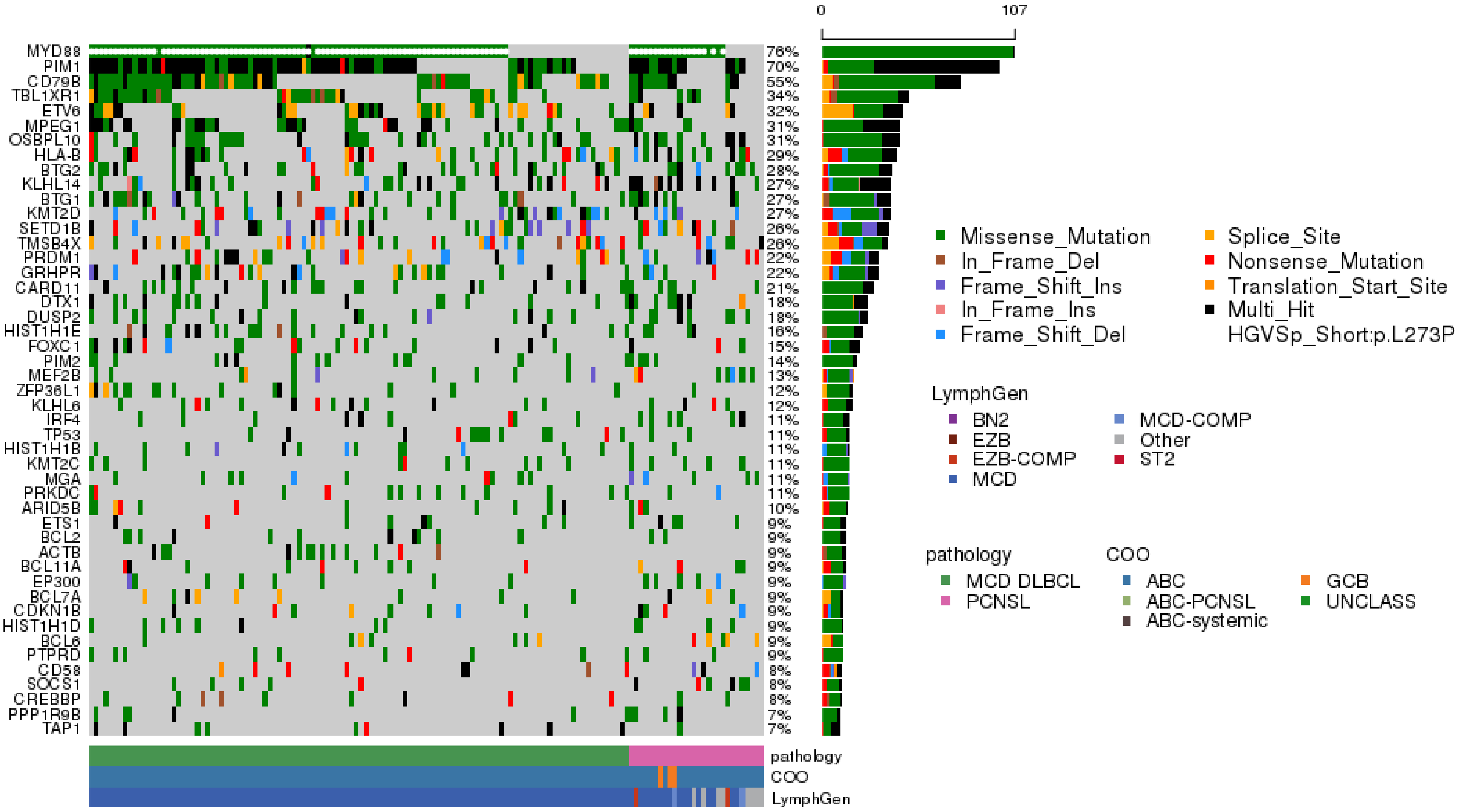


**Supplemental figure 9**

PCNSL vs. (MCD) ABC-DLBCL exomes (Schmitz, Chapuy, Reddy)

### Unique mutations in PCNSL compared to systemic MCD ABC-DLBCL

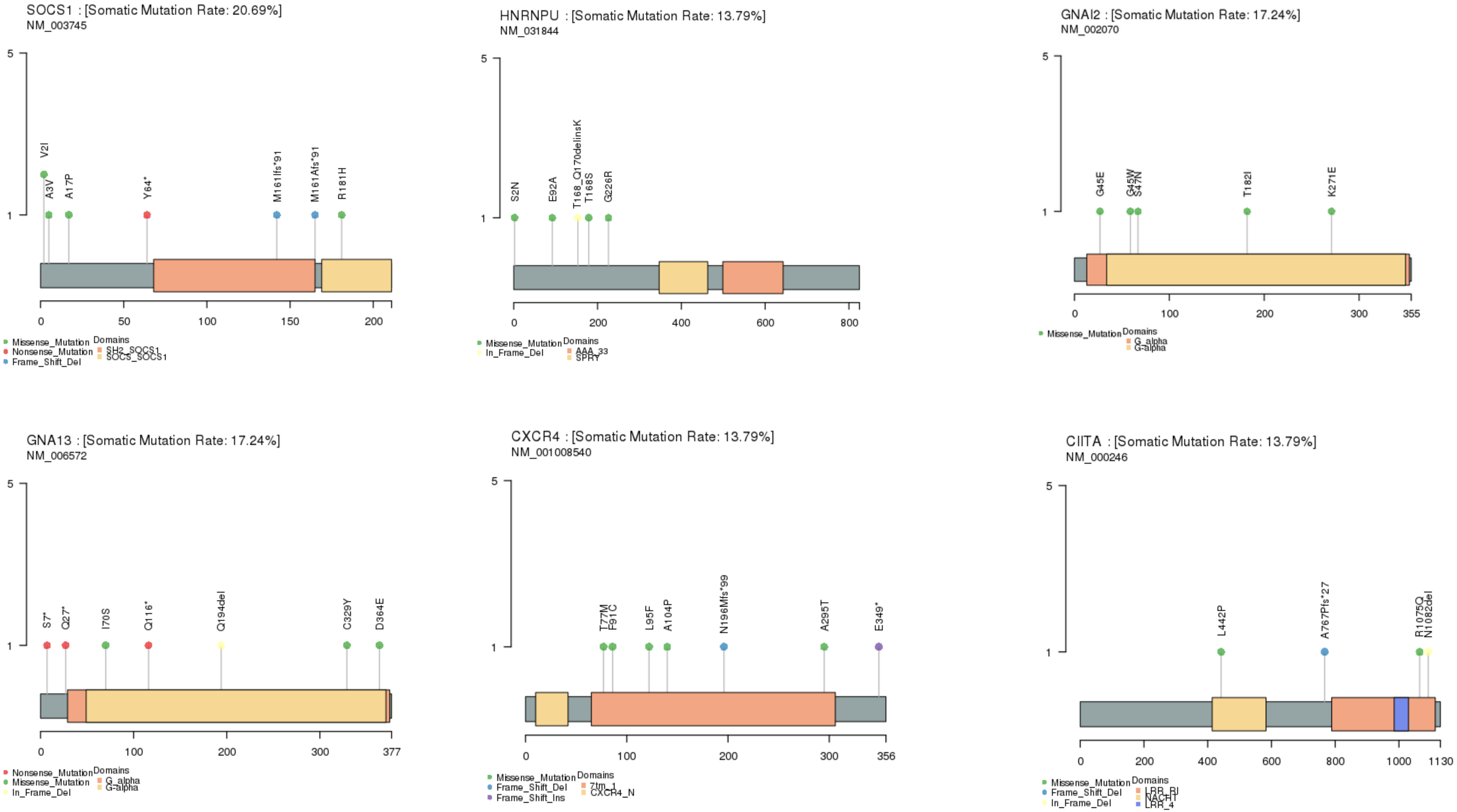


**Supplemental figure 10**
