## supplemental figures 1-10 legends for "Primary Central Nervous System Lymphoma Tumor Biopsies Show Heterogeneity in Gene Expression Profiles and Genetic Subtypes"

Supplemental figure 1. **Reproducibility of nCounter data over time.** Graphs show the correlation of gene expression data (raw and normalized) obtained from a DLBCL cell line (DB) pelleted and embedded in a paraffin block run 1 year apart using the nCounter Pan Cancer panel. The resulting data were highly correlated using the Pearson correlation test indicating the stability of results from this method.

Supplemental figure 2. **Immunohistochemistry staining for FGF1 and FGFR2** (all photomicrographs at 40X).

Top row, left to right: hematoxylin and eosin (H&E), negative control, FGF1, FGFR2 of kidney. Photomicrographs demonstrate FGF1 protein expression glomerulus in contrast to FGFR2 staining in tubules, demonstrating specificity of staining.

Second row, left to right: H&E, negative control, FGF1, FGFR2 of normal lymphoid tissue in a tonsil. Photomicrographs demonstrate very little staining of lymphocytes or other cells.

Third row, left to right: H&E, negative control, FGF1, FGFR2 of normal brain. Photomicrographs demonstrate little to no staining.

Fourth row, left to right: H&E, negative control, FGF1, FGFR2 of example PCNSL case. Photomicrographs demonstrate FGF1 staining in area of malignant lymphoma cells while FGFR2 is negative.

Fifth row, left to right: H&E, negative control, FGF1, FGFR2 of brain parenchyma adjacent to PCNSL tumor cells. Photomicrographs demonstrate a few FGF1 positive cells and FGFR2 staining as well.

Supplemental figure 3. **Distribution of mutations across the aSHM sites.** Scatterplot showing the distribution of mutations for each sample across regions of aSHM, separated by gene.

Supplemental figure 4. **Oncoplot and table of PCNSL separated by transcriptional subtype, cell of origin, and LymphGen genetic classification.** No genetic lesions were enriched in either the P1 or P2 groups; nor were genetic subtypes based on LymphGen associated with P1 or P2 transcriptional groups.

Supplemental figure 5. **Fraction of EBV reads in PCNSL exomes**. Boxplot showing the proportion of reads in the PCNSL WES libraries that align to the EBV genome according to GATK PathSeq. As a comparison, WES libraries from lymphoma that have been verified for EBV status by EBER *in-situ* hybridization are included as comparison. All the PCNSL samples showed 0 reads aligning to the EBV genome, except one case, which showed some read support for EBV, though it was just below the threshold for consideration (0.0002%).

Supplemental figure 6. **Plot showing GISTIC amplification and deletion peaks in PCNSL samples (n=16).** Only peaks that showed enrichment in a gene from a curated list of lymphoma-related genes were annotated. Only samples that had a purity>0.4 as determined by PureCN were included in CNV analysis.

Supplemental figure 7. **Mutational profile of ABC cell lines.** The genetic subtypes of the five cell lines were analyzed using LymphGen algorithm. The three ABC-DLBCL cell lines (TMD-8, OCILY3, HBL-1) had mutations that are canonically associated with the MCD subgroup, while the two ABC-PCNSL cell lines (TK_cell_line, HKBML) were classified as Other.

Supplemental figure 8. **Mutational landscape of PCNSL versus nodal ABC-DLBCL from published sources.** Oncoplot showing the mutational landscape of nodal ABC-DLBCL from WES (left) compared to the mutational landscape of PCNSL (right).

Supplemental figure 9**. Mutational landscape of PCNSL versus nodal MCD-ABC-DLBCL from published sources.** Oncoplot showing the mutational landscape of nodal ABC-DLBCL from WES, stratified to those that were classified as MCD by the LymphGen algorithm (left) compared to the mutational landscape of PCNSL (right).

Supplemental figure 10. **Lollipop plots highlighting unique mutations in PCNSL compared to systemic MCD ABC-DLBCL.** The distribution of mutations in *SOCS1*, *HNRNPU*, *GNAI2*, *GNA13, CXCR4,* and *CIITA* in PCNSL.
